# Investigating the genetic factors of medication dosing using biobank-linked drug purchase data

**DOI:** 10.1101/2025.04.01.25325019

**Authors:** Silva Kasela, Laura Birgit Luitva, Kristi Krebs, Estonian Biobank Research Team, Märt Möls, Lili Milani, Maris Alver

## Abstract

Despite significant pharmacogenomic (PGx) insights into key biological pathways influencing drug response, the polygenic contribution to dose variability and the potential of electronic health records (EHRs) for maintenance dose estimation remain largely unexplored. To address this gap, we leveraged longitudinal drug purchase data from the Estonian Biobank to derive medication dose metrics and investigate genetic associations using polygenic scores (PGS) and genome-wide association studies (GWAS). Daily doses per purchase as well as median and maximum doses as consolidated metrics across purchases were derived for statins, warfarin, metoprolol, antidepressants, and antipsychotics. Sample sizes ranged from 684 (antipsychotics) to 20,642 (statins), with median doses reflecting typical maintenance doses. The PGS for the indicator trait was significant for the daily dose of statins (coronary heart disease PGS, β = 0.02, P = 5.9×10^− 10^) and metoprolol (systolic blood pressure PGS, β = 0.03, P = 7.5×10^−13^). The PGS for body mass index was linked to daily doses of statins (β = 0.02, P = 6.4×10^−7^), metoprolol (β = 0.03, P = 1.4×10^−14^), and warfarin (β = 0.03, P = 0.001), whereas the PGS for educational attainment showed opposing associations with statins (β = –0.01, P = 5.9×10^−4^) and antidepressants (β = 0.01, P = 0.002). Median and maximum doses had similar but weaker associations. GWAS confirmed signals for metoprolol (*CYP2D6*, P = 1.1×10^−20^) and warfarin (*CYP2C9*, P = 8.9×10^−60^; *VKORC1*, P = 4.2×10^−148^), as well as enrichment of PGx signals for individual statins (P = 0.02 for simvastatin, P = 0.03 for atorvastatin). Altogether, these findings illustrate the feasibility and value of deriving medication doses from EHRs and highlight the role of polygenic liability and PGx factors in dose variability.

## BACKGROUND

Understanding individual variability in drug response is a cornerstone of precision medicine. While drug response is influenced by a combination of genetic, environmental, and lifestyle factors, pharmacogenomics (PGx) has provided the most significant insights into medication metabolism and efficacy^1^. Much of the progress has focused on genetic variation in key pathways influencing pharmacokinetics, such as cytochrome P450 enzymes, which drive significant interindividual variability in drug metabolism and, consequently, impacts drug efficacy, dosing, and safety^2,3^. Similarly, genetic variation in drug transporters, drug targets, and absorption, distribution, and excretion pathways has been shown to play an equally critical role in shaping the pharmacokinetics and pharmacodynamics of medications. However, research in this area has been limited by a narrow focus on single genes, leaving the broader genetic contribution of polygenic traits largely unexplored^4–7^.

The advent of large-scale biobanks linked with longitudinal electronic health records (EHRs) has transformed biomedical and genomics research by enabling access to real-world health data at an unprecedented scale. These resources provide unparalleled power and feasibility for exploring research questions previously impeded by limited sample sizes and data scarcity. For instance, such resources have contributed to the discovery of genetic variants associated with complex traits, facilitated the translation of these findings into improved understanding of disease mechanisms and therapeutic targets, and enabled novel method development^8–10^. While much of the research focus has been on pooling disease endpoints, biomarkers, and easily accessible traits, there is growing interest in using real-world health data to map disease networks^11–13^, track longitudinal biomarker trends^14–16^, and assess treatment outcomes^17–19^.

Genomic research utilizing treatment data from EHRs has primarily examined metrics such as medication use (yes/no), counts and types of medications, treatment stability, and discontinuation, with results often reflecting underlying disease associations. For instance, genome-wide association studies (GWAS) on medication use data^17,18^ largely identified genetic loci linked to conditions for which the medications were prescribed, indicating that the genetic factors influencing medication use overlap with those driving disease risk, as expected. By contrast, investigations into the genetic basis of treatment response using EHRs have revealed few or no genome-wide significant hits^20–24^ and low genetic heritability^5,19,24,25^, suggesting that the genetic effects on treatment response may be modest^24^, mitigated by confounding factors such as disease severity or treatment^19^, and difficult to detect with current approaches. While numerous methods have been developed for inferring medication use periods from longitudinal EHRs, with data-driven approaches recognized for providing more accurate estimates^26^, methodologies for calculating daily dose values using EHRs are scarce^27,28^, and genome-wide assessments of medication doses have largely remained unexplored.

In this study, we leveraged longitudinal drug purchase data from the Estonian Biobank (EstBB)^29^ to derive daily dose values, capturing real-world treatment adjustments and medication use over time. By linking the dose estimates, derived across a range of medications, to genetic data, we aimed to investigate their association with the polygenic liability to complex traits as well as conduct genome-wide scans to explore genetic contributions to medication dose variability.

## METHODS

### The Estonian Biobank (EstBB)

EstBB is a volunteer-based population biobank in Northern Europe with continually updated national health registry linkages and genotype data for 212,000 individuals. The activities of EstBB are regulated by the Human Genes Research Act, adopted in 2000 specifically for the operations of EstBB. All EstBB participants have signed a broad informed consent. The current study was approved by the Estonian Committee on Bioethics and Human Research at the Estonian Ministry of Social Affairs (24 March 2020, nr 1.1-12/624) and carried out using data according to release U16 from EstBB. Data freeze 2024v1 with follow-up data until 16/04/2024 at EstBB was used for the analyses. Individuals of European ancestry (>99.7% of the cohort) were considered in all analyses^29^.

### Treatment variables

We focused on two main medication groups: cardiovascular drugs, including statins (ATC code C10AA*, and their milligram content in combination drugs, e.g., C10BA*, C10BX* that were grouped with their respective single-drug statins), warfarin (B01AA03), and metoprolol (C07AB02); and psychiatric drugs, including antidepressants (N06A*) and antipsychotics (N05A*, except lithium). Doses were analysed both across drug classes and for individual medications within each class. For individual medications within each class, we considered time periods when only the drug of interest was purchased relative to all other drugs in the medication class, to exclude time periods of simultaneous use of multiple drugs from the same class. To account for variability in the data (e.g., trial periods with another drug), we focused on time periods during which the drug of interest accounted for at least 75% of purchases over all purchases in the medication class, using a 1-year sliding window from each purchase date. These criteria were applied across the following medication classes with sufficient sample size to identify individual drug use: C10AA* and C10B* for statins, N06A* for antidepressants, C07* for metoprolol, and B01* for warfarin. The individual medications considered for each drug class are outlined in **Supplementary Table 1**. Warfarin and metoprolol were selected for their narrow therapeutic range and strong established PGx associations.

### Sample selection

Individuals were required to have purchase records spanning more than 365 days, either within a medication class or for a specific drug, with at least six purchases on unique dates, to exclude those with short-term use. Purchases made before age 18 were excluded. For antidepressants and antipsychotics, we included individuals whose main ICD-10 code on the prescription was depression (ICD-10 F32, F33, F41.2) and schizophrenia spectrum disorder (ICD-10 F20-F29), respectively, to exclude variability introduced by lower doses prescribed for other indications. This criterion was not applied to statins, as dosing for statins is primarily driven by cardiovascular risk and lipid levels, with minimal variability stemming from alternative clinical uses.

We required a minimum of 500 individuals per medication after applying all filtering criteria, which limited the analyses to a subset of drugs within each class. Simvastatin, atorvastatin, and rosuvastatin were included among statins, while escitalopram, sertraline and fluoxetine passed this threshold among antidepressants. No individual antipsychotic drugs surpassed the minimum sample size requirement.

### Derivation of medication dose variables

To derive dose variables, we used digital drug dispensing data from the Estonian Health Insurance Fund (available since 2004) linked with EstBB. The digital drug dispensing data include all medications prescribed in the healthcare system. The dataset contains information on drug prescription and purchase dates, the dose and content per package, and the number of packages bought. We derived two dose variables for each drug class and three dose variables for each individual medication. First, to capture the longitudinal nature of drug dispensing data and account for sample-specific drug patterns over time, we considered all individual purchases and calculated daily doses from each purchase per sample. Second, as a consolidated metric, we calculated a median dose for each individual, either across all purchases within a medication class or for a specific medication. Third, we determined a maximum dose variable for individual medications, which reflects peak treatment intensities and are independent of the aggregation of purchases into a single variable (**Supplementary Figure 1**). For analyses, a binary maximum dose variable was determined based on whether the individual had purchased the medication with the highest dose or not.

Specifically, the daily dose per purchase was calculated by multiplying the package content (dose in milligrams and number of pills) by the number of packages bought. Purchases within two weeks were consolidated with the purchase date assigned to the earliest date within the 14-day window. Next, the calculated *mg x pills in a package x number of packages* variable for each purchase was divided by the number of days until the next purchase, resulting in an estimate of a daily dose per purchase. Purchases separated by more than 365 days were treated as distinct purchase batches, and daily doses per purchase were calculated within these batches. The last purchase per batch served as an end date for the preceding purchase only. Batches consisting of only a single purchase were excluded. When pooling statins, antipsychotics, and antidepressants to assess class-level associations, daily doses were standardized using the Defined Daily Dose (DDD) factors as outlined by WHO^30^. For antipsychotics, long-acting injectables were first converted to daily doses based on the minimum duration recommendation according to the drug-specific treatment guidelines.

The median dose variable was calculated by taking the median of all derived daily doses. If an individual had breaks in treatment (i.e., >365 days between purchases) and daily doses per purchase were calculated within batches, the median dose was first calculated within each batch, followed by a subsequent median calculated across all batches.

For the maximum dose variable, we considered the highest dose (mg on package) purchased at least three times based on unique purchase dates (not consolidated). Binary variables for maximum dose were defined by contrasting the highest dose in milligrams with all other doses. In case the highest dose was available for less than 100 individuals, it was combined with the next highest dose level to ensure sufficient representation. The counts for maximum dose are outlined in **Supplementary Table 2**.

### Treatment length

Given that treatment duration correlates with increases in medication dose and thus affects the derivation of both median and maximum doses, we calculated the length of treatment for each individual. For this, we summed the days’ supply (i.e., the total number of pills) for each purchase and converted the total into years, assuming the intake of one pill per day (**Supplementary Figure 1**). While the frequency of purchases varies across individuals, this approach standardizes treatment duration based on the total days’ supply, independent of purchase events.

### Genotype data

All EstBB participants were genotyped using Illumina GSAv1.0 and GSAv2.0 arrays, as well as GSAv2.0_EST and GSAv3.0_EST arrays with add-on SNV content from EstBB whole genome sequencing data, aligned to the GRCh37 reference genome. Quality control was conducted according to best practices: exclusion of individuals with call rate <95%, with mismatch between genotype and phenotype sex or who deviatedJ±3SD from the samples’ heterozygosity rate mean; exclusion of SNVs with call rate <95%, HWE p<1e-4, deemed ambiguous (A/T and C/G) or invariable, showing potential traces of batch bias, or poor cluster separation and inconsistent allele frequency among any of the EstBB genotyping experiments^31,32^. Pre-phasing was carried out with Eagle v2.3^33^ and imputation with Beagle^34^ v.28Sep18.79367 using the Estonian population-specific reference panel built based on 2,695 whole genome sequencing samples^35^. The X chromosome was processed independently. The pseudoautosomal (PAR) and non-PAR regions were phased separately. For males, any heterozygous calls in non-PAR region were excluded.

### Polygenic scores

For polygenic score (PGS) calculation, we used GWAS summary statistics based on GRCh37 for different complex traits relevant to the medications of interest. European-specific results were preferred when available. To evaluate the suitability of these traits for PGS calculation and assess genetic relationships between them, we examined SNP-based heritability to ensure estimates were not deflated due to low power in the underlying GWAS (**Supplementary Figure 2A**) and analyzed pairwise genetic correlation using LDSC^36^ v1.0.1 (**Supplementary Figure 2B**). GWAS summary statistics were restricted to SNVs in the HapMap 3 reference panel^37^ with LD scores derived from the European ancestry samples. The results were visualized using R/*corrplot*^38^ and the hierarchical clustering method.

PGSs were calculated with PGS-CS^39^, a Bayesian polygenic prediction method that applies a continuous shrinkage prior on SNV effect sizes and infers posterior SNV weights using GWAS summary statistics restricted to 1.1 million HapMap variants^37^ and an external European sample-based LD reference from the 1000 Genomes Project^40^. EstBB genotype data were pre-filtered for MAF ≥0.1% and imputation quality ≥0.8. The default parameters and the *auto* option were used, and the HLA region (6:25000000-35000000) was excluded. For schizophrenia (SCZ) and major depressive disorder (MDD), the HLA region was included, given the significant association of the region in respective GWAS. Additionally, the global shrinkage parameter phi = 1 was used for SCZ, to capture the highly polygenic architecture of the disorder^41–45^. Pearson correlation was used to assess pairwise PGS correlations and results were visualized with R/*corrplot*^38^ (**Supplementary Figure 2B)**.

Based on SNP-based heritability and correlation analyses among underlying GWAS and calculated PGSs, which confirmed expected genetic relationships and consistency in estimates, we included the following eleven PGSs for all medication dose analyses: educational attainment (EA)^46^, ever smoking (EVERSMK)^47^, estimated glomerular filtration rate (eGFR)^48^, alanine aminotransferase (ALT)^49^, body mass index (BMI)^50^, glycated haemoglobin (Hb1Ac)^51^, LDL-cholesterol (LDL-C)^52^, HDL-cholesterol (HDL-C)^52^, triglycerides (TG)^52^, and systolic blood pressure (SBP)^53^. We additionally included the following PGSs: i) SCZ^41^ and treatment-resistant SCZ^54^ for antipsychotics; ii) MDD^55^ and antidepressant response^56^ for antidepressants; iii) coronary heart disease (CHD)^57^ for statins and metoprolol; and iv) CHD and venous thromboembolism (VTE)^58^ for warfarin. Bonferroni correction was applied based on the number of PGSs considered for each drug class or drug, which resulted in the following thresholds: 0.05/11 = 0.0045 for statins and metoprolol, and 0.05/12 = 0.004 for antidepressants, antipsychotics, and warfarin.

We excluded individuals who were included in the GWAS analyses used for calculating PGSs in the current study and one member per related pairs (PLINK PI_HAT >0.2) when testing the association between PGSs and derived dose variables^31,32^. PGSs were standardized to Z-scores with a mean of 0 and a standard deviation (SD) of 1 across all EstBB individuals, with subsets extracted for specific analyses.

### Association testing with medication dose variables

#### Longitudinal assessment of individual daily doses

To capture the longitudinal nature of drug dispensing data and account for sample-specific drug use patterns over time, we employed linear mixed modelling (R/*lme4*, R/*lmerTest*^59,60^) to test the association between derived daily doses and PGSs. We restricted the analysis to individuals whose first purchase of the drug class or specific drug of interest occurred on or after 01/01/2005, allowing us to capture all consecutive purchases from the start of treatment while considering that electronic health records in EstBB became available in 2004. A Box-Cox transformation was used to determine the optimal transformation of the response variable, leading to a log-scale transformation. Daily doses greater than 3SD from the mean on a log scale were excluded. Fixed effects included population-level parameters (sex, age at each purchase, years since the first purchase, and ten principal components (PCs) of the genotype data), while subject-specific effects (subject-specific random intercept and subject-specific effect of time from the first purchase) were considered as random effects. Normality assumption for the daily dose variable was assessed with diagnostic plots of model residuals, and the BOBYQA optimizer^61^ was applied in all analyses. To ascertain independent PGSs associated with daily doses, we applied linear mixed modelling using a forward stepwise approach. Namely, each PGS of interest for the considered variable was first modelled individually with the daily dose as a dependent variable. Next, the PGSs were incrementally added to the main model, starting with the one with the lowest p-value and scanning through all other PGSs to determine independent associations. The process was repeated until no additional PGSs were significant at Bonferroni-corrected threshold specific to the medication under investigation.

As a sensitivity analysis for statins, we additionally included a binary variable indicating whether the purchase occurred before or after a hard cardiovascular event (myocardial infarction or cerebral infarction, ICD-10 codes I21, I22, I63), as statin doses are commonly increased following these events to manage heightened cardiovascular risk. Among 20,642 statin users, 2,395 simvastatin users, 9,400 atorvastatin users, and 11,279 rosuvastatin users, 3,440 (17%), 531 (22%), 2,214 (24%), and 1,466 (13%) experienced an event during treatment, respectively.

#### Association testing with median dose and maximum dose

To test the association between the median dose derived across all purchases with PGSs, we applied multivariate linear regression using a forward stepwise approach, as previously outlined, with sex, birth year, treatment length on supply and the first ten genotype PCs as covariates. A Box-Cox transformation was used to determine the optimal transformation of the response variable, resulting in a log-scale transformation. Individuals with a median dose exceeding 3SD from the mean on a log scale were excluded, as were those with a treatment length of less than two years (based on days’ supply) due to the high variability in median dose values observed for shorter treatment durations (**Supplementary Figure 3**). Logistic regression in a forward stepwise manner was similarly used for the binary maximum dose variable with sex, birth year, treatment length on supply and the first ten genotype PCs as covariates.

#### Genome-wide association studies

GWAS were performed using REGENIE^62^ v3.4, employing a 2-step regression model (additive test) for quantitative traits (median doses) and binary traits (maximum doses), which incorporates a genetic relationship matrix, hence no related samples were excluded. SNVs with MAF <0.5% and info score <0.8 were excluded. One individual from each monozygotic twin pair was excluded. Individuals whose median dose was greater than 3SD from the mean on a log scale and whose treatment length was less than two years (based on days’ supply) were excluded. Rank-based inverse normal transformation was applied to each median dose variable. Two GWAS analyses were performed for the same medication dose variable: the first included sex, birth year, treatment length, and the first ten genotype PCs as covariates; the second additionally incorporated the PGS for the primary phenotypic trait associated with the drug (i.e., CHD PGS for statins, SCZ PGS for antipsychotics, MDD PGS for antidepressants, SBP PGS for metoprolol, and VTE PGS for warfarin) to search for effects beyond the underlying genetics of the disease. Individuals included in the GWAS analyses used for calculating PGSs were excluded in the latter analyses. Regional association plots for the significant loci were generated using LocusZoom^63^.

#### PGx gene associations in GWAS

To determine whether known PGx genes were captured among top GWAS results, despite not reaching genome-wide significance, we first assessed whether genetic variants within these genes collectively showed enrichment of association signal and additionally identified which drug-relevant PGx genes showed the strongest associations. To this end, we compiled gene sets specific to each medication and medication class. These included genes encoding pharmacokinetically relevant proteins involved in the absorption, distribution, metabolism, and excretion (ADME) of drugs, as well as all genes highlighted in the published guidelines of the Clinical Pharmacogenetics Implementation Consortium (CPIC) (**Supplementary Table 3**). Gene lists were obtained from the Pharmacogenomics Knowledge Base (PharmGKB)^64,65^ by downloading all pharmacokinetic pathway tables for the drugs and drug classes analyzed in this study (accessed November 5, 2024).

To map genetic variants to genes, SNVs were assigned to protein-coding genes using BEDtools v2.31.0^66^ and GENCODE gene coordinates (GRCh37), including ±5 kb flanking regions. For each gene, the SNV with the lowest p-value was selected (i.e., retaining one SNV per gene), and those corresponding to the PGx gene set of the medication of interest were extracted. To account for LD, SNVs in the PGx gene set were pruned using a 50-kb window, a step size of 5 SNVs, and an r^2^ threshold of 0.2. A background set was defined by considering the top SNVs for all protein-coding genes, including PGx genes, which were LD-pruned with the same parameters. Analyses were conducted for drugs with pruned PGx gene sets containing at least ten genes, namely statins, simvastatin, atorvastatin, rosuvastatin, antidepressants, sertraline, and antipsychotics.

To test whether medication-specific PGx genes showed stronger associations than expected by chance in the respective GWAS results, we compared the median p-value of selected SNVs in the PGx gene set to an empirical null distribution generated by permutations. Median p-value was chosen to represent the overall signal of PGx genes while reducing the influence of outliers. A lower median p-value for SNVs in the PGx gene set would indicate an overall enrichment of association signals compared to the null. Specifically, we compared the median p-value of SNVs in each medication-specific PGx gene set to a null distribution obtained by randomly sampling an equal number of SNVs from a background set of pruned SNVs across all protein-coding genes over 10,000 iterations. The permutation p-value was calculated as the proportion of permuted median p-values that were as low as or lower than the observed median p-value.

To identify PGx genes with the strongest associations with dose variables, we applied an empirical threshold-based approach. The significance threshold was defined as the 5^th^ percentile of p-values from an LD-pruned background set of SNVs for each medication or medication class. SNVs within the corresponding LD-pruned PGx gene set with p-values below this threshold were considered significant.

Statistical analyses were conducted with R software version 4.4.2^67^ .

## RESULTS

### Study overview and sample characteristics

To test the association of genetic factors with medication doses, we considered three drug classes: statins, antidepressants and antipsychotics. We also focused on individual medications within these classes that had at least 500 individuals meeting the filtering criteria, as well as included metoprolol and warfarin (**Figure 1**). The sample sizes ranged from 684 to 20,642 with fluoxetine having the lowest mean number of purchases (mean=15.4, SE=13) and antipsychotics the highest (mean=41.4, SE=32.3). All medications had higher purchase rates among females, ranging from 85.3% for fluoxetine to 51.5% for warfarin. Antipsychotics and antidepressants had the lowest age at first purchase (38.8 and 39.8 years, respectively) compared to statins, metoprolol and warfarin (>58 years). The time between the first and last purchase ranged from 4.1 years for sertraline to 9.1 years for antipsychotics (**Table 1**). As expected, the most common prescription diagnoses were hypercholesterolemia (ICD-10 E78) for statins, depression (ICD-10 F32, F33, F41.2) and other anxiety disorders (ICD-10 F41) for antidepressants, atrial fibrillation (ICD-10 I48) for warfarin, and hypertension (ICD-10 I10-I11) for metoprolol. For antipsychotics, however, other anxiety disorders (ICD-10 F41) and sleep disorders (ICD-10 G47) were the most prevalent (**Supplementary Figure 4**). The distribution of daily doses per purchase for each drug class and individual medications by the indicated diagnosis on the prescription is outlined in **Supplementary Figure 5**.

**Figure 1.**
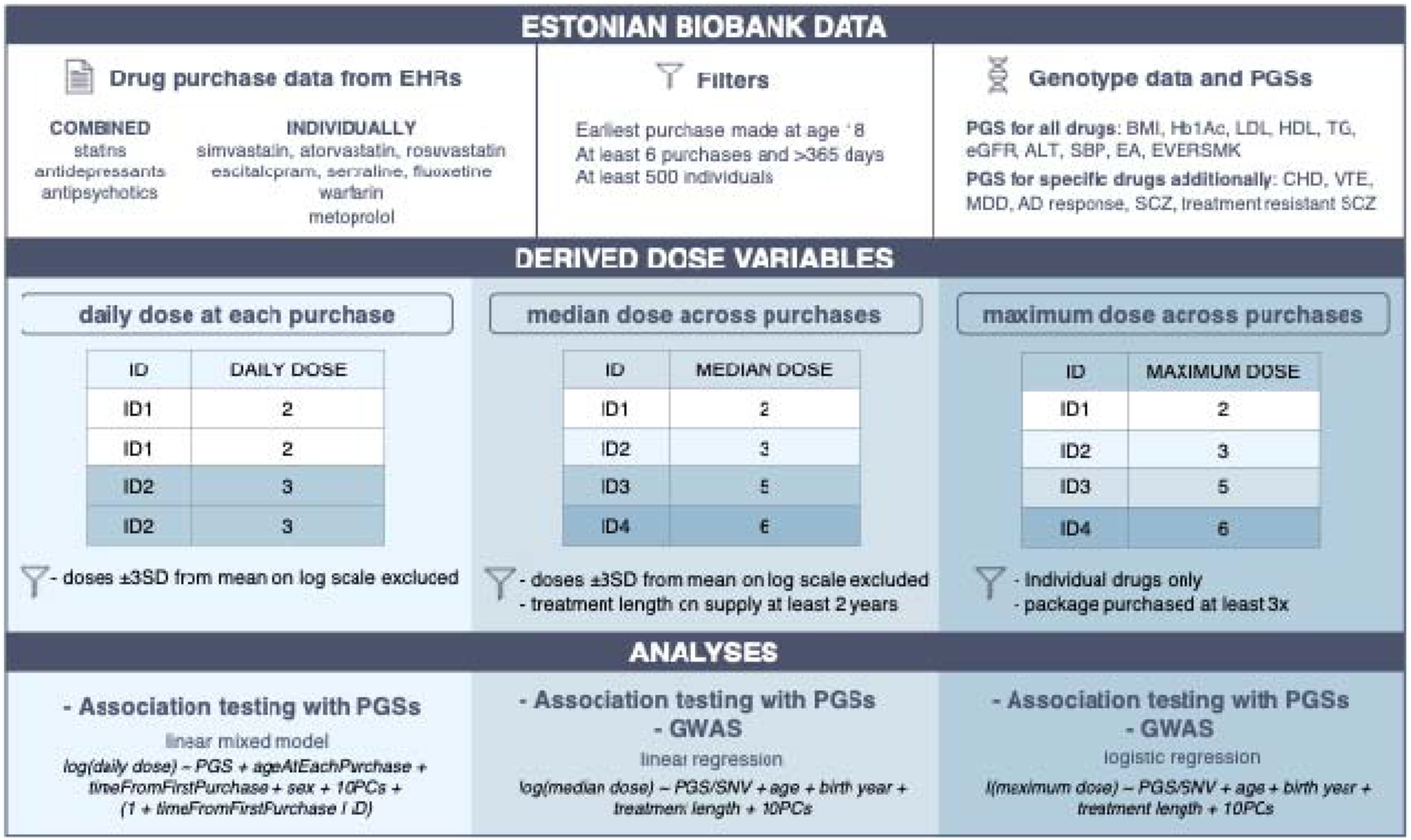
Overview of the study design. Purchase data were used to derive three dose variables: (i) daily dose at each purchase, (ii) median dose across purchases, and (iii) maximum dose across purchases. PGSs were tested for associations with the dose variables using linear mixed models for daily doses, linear regression for median doses and logistic regression for maximum doses, with GWAS performed for the latter two measures. I(max dose) is the indicator function that is 1 if the person has the max dose, and 0 otherwise. BMI – body mass index, HbA1c – glycated haemoglobin, LDL – low-density lipoprotein, HDL – high-density lipoprotein, TG – triglycerides, eGFR – estimated glomerular filtration rate, ALT – alanine aminotransferase, SBP – systolic blood pressure, EA – educational attainment, EVERSMK – ever smoking, CHD – coronary heart disease, MDD – major depressive disorder, SCZ – schizophrenia, VTE – venous thromboembolism, AD – antidepressant.

**Table 1.** Overview of the study cohort.

| Drug | N | N of purchases on unique dates (mean, SD) | Female (%) | Age at first purchase (mean, SD) | Time from first to last purchase in years (mean, SD) |
| --- | --- | --- | --- | --- | --- |
| **STATINS** | 20,642 | 27.0 (22.3) | 12,908 (62.5%) | 59.7 (10.4) | 7.3 (5.1) |
| Simvastatin | 2,395 | 30.1 (22.5) | 1,578 (65.9%) | 60.7 (10.0) | 7.4 (4.9) |
| Atorvastatin | 9,400 | 23.4 (19.0) | 5,555 (59.1%) | 62.0 (10.6) | 5.7 (4.2) |
| Rosuvastatin | 11,279 | 24.5 (21.0) | 7,254 (64.3%) | 60.1 (10.4) | 6.5 (4.9) |
| **ANTIDEPRESSANTS** | 9,877 | 21.3 (21.4) | 7,545 (76.4%) | 39.8 (14.4) | 6.7 (5.2) |
| Escitalopram | 3,392 | 17.0 (15.5) | 2,727 (80.4%) | 41.8 (14.1) | 5.1 (4.4) |
| Sertraline | 1,379 | 17.2 (16.4) | 1,137 (82.5%) | 40.22 (15.2) | 4.1 (3.9) |
| Fluoxetine | 919 | 15.4 (13.0) | 784 (85.3%) | 41.3 (13.7) | 4.7 (4.2) |
| **ANTIPSYCHOTICS** | 684 | 41.4 (32.3) | 438 (64.0%) | 38.8 (15.4) | 9.1 (5.5) |
| **METOPROLOL** | 15,254 | 31.5 (25.0) | 10,255 (67.2%) | 58.5 (12.8) | 7.8 (5.4) |
| **WARFARIN** | 1,839 | 27.8 (19.7) | 948 (51.5%) | 63.2 (12.2) | 6.0 (4.0) |

### Association of derived daily doses with polygenic scores

To investigate the associations between PGSs and daily doses, we applied linear mixed modelling. Each PGS was first tested individually, and independent associations were identified using a forward stepwise approach by incrementally adjusting for additional PGSs. The analyses revealed that both CHD PGS and BMI PGS were consistently and independently associated with higher daily doses across all statin medications, observed both at the class level (CHD PGS: β=0.02, SE=0.004, P=5.9×10^−10^; BMI PGS: β=0.02, SE=0.004, P=6.4×10^−7^) and for individual drugs: simvastatin (CHD PGS: β=0.03, SE=0.009, P=7.4×10^−4^; BMI PGS: β=0.03, SE=0.008, P=1.5×10^−4^), atorvastatin (CHD PGS: β=0.04, SE=0.005, P=5.4×10^−13^; BMI PGS: β=0.02, SE=0.005, P=7.4×10^−4^), and rosuvastatin (CHD PGS: β=0.03, SE=0.004, P=9.4×10^−13^; BMI PGS: β=0.02, SE=0.004, P=2.7×10^−7^) (**Figure 2A**). Interestingly, among Bonferroni-significant associations observed when PGSs were tested individually, EA PGS stood out, showing a significant negative correlation with daily doses for all statins (β=–0.01, SE=0.004, P=5.9×10^−4^) and specifically for atorvastatin (β=–0.02, SE=0.005, P=6.3×10^−4^) and rosuvastatin (β=–0.02, SE=0.004, P=2.2×10^−4^) (**Supplementary Table 4**). However, these associations were no longer significant in forward regression models, suggesting that its genetic liability was already accounted for by CHD PGS and BMI PGS. Including a hard cardiovascular event as a covariate slightly attenuated the associations with CHD PGS and BMI PGS, but their effects remained significant (**Supplementary Figure 6**). This indicates that while such events typically prompt more intensive therapy, genetic predisposition to cardiovascular disease and body composition still contribute significantly to increased statin dose.

**Figure 2.**
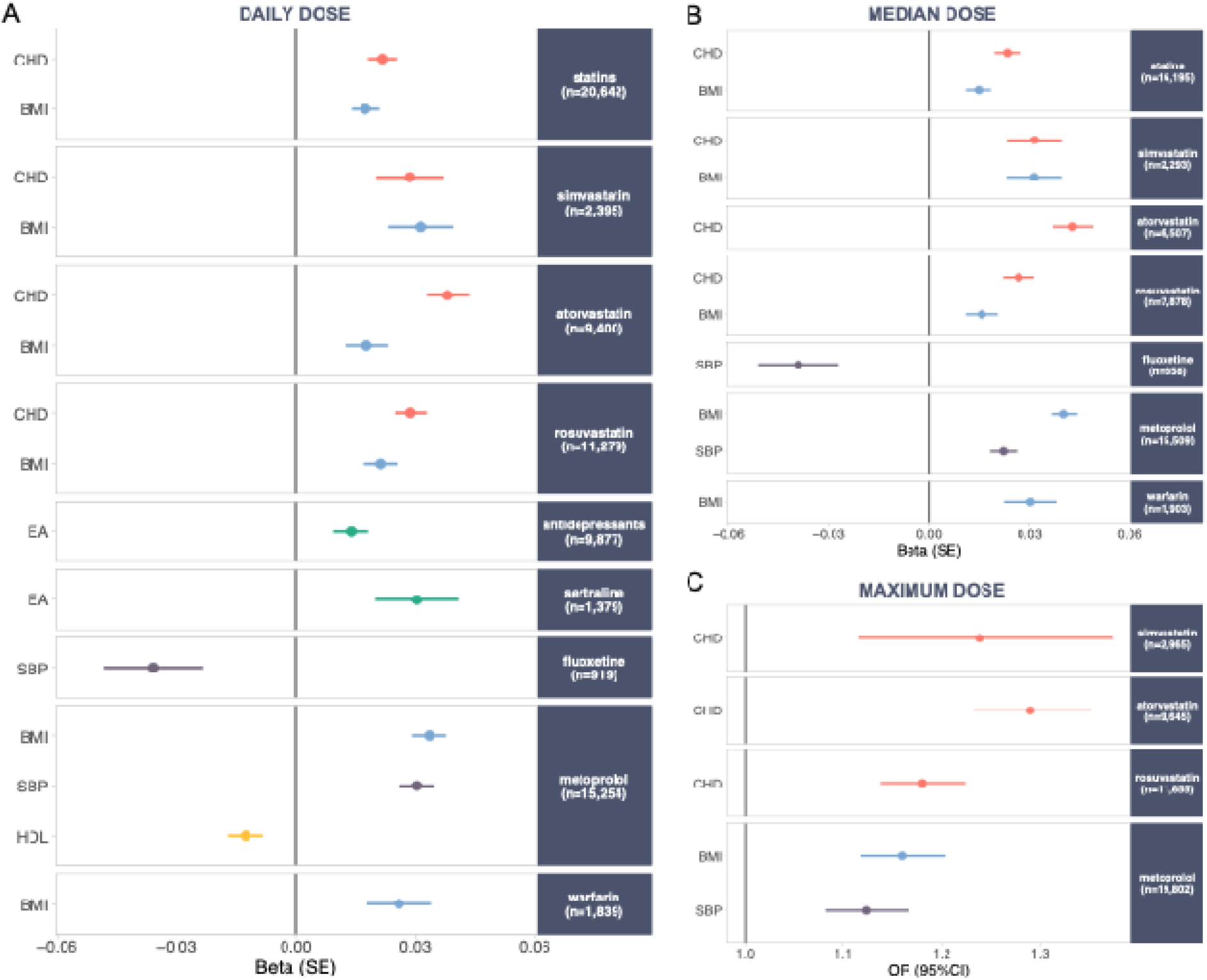
Effect sizes for PGSs significantly associated with derived dose variables. Only PGSs that surpassed Bonferroni correction and were independently associated are shown for (A) daily doses, (B) median doses, and (C) maximum doses. Different colours represent distinct PGSs: red for coronary heart disease (CHD), blue for body mass index (BMI), green for educational attainment (EA), purple for systolic blood pressure (SBP), and yellow for high-density lipoprotein (HDL).

Among individuals taking antidepressants, EA PGS showed a significant positive correlation with daily doses overall (β=0.01, SE=0.004, P=0.002) and specifically for sertraline (β=0.03, SE=0.01, P=0.004). In contrast, SBP PGS was negatively associated with the daily doses of fluoxetine (β=–0.04, SE=0.01, P=0.004). Among those taking metoprolol, BMI PGS (β=0.03, SE=0.004, P=1.4×10^−14^), SBP PGS (β=0.03, SE=0.004, P=7.5×10^−13^), and HDL PGS (β=–0.01, SE=0.004, P=0.004) were all independently linked with the daily doses, whereas among warfarin users, only BMI PGS (β=0.03, SE=0.008, P=0.001) showed an association (**Figure 2A**). No significant associations were observed for antipsychotic users or individuals taking escitalopram, nor did trait-specific PGSs (SCZ, MDD, VTE) show nominal correlations for any antidepressants, antipsychotics and warfarin, respectively (**Supplementary Table 4**).

### Association of median and maximum doses with polygenic scores

To provide consolidated measures of typical maintenance and peak dosing levels, we derived two summary dose metrics for each drug: the median dose, calculated across all purchases, and the maximum dose, defined as the highest dose purchased at least three times (**Figure 1**). While the derived median doses reflected lower-bound estimates of typical maintenance doses (**Supplementary Table 5**), lower doses were observed for antipsychotics, and simvastatin (**Table 2**). Consistent with trends seen for daily doses per purchase (**Supplementary Figure 5**), the distribution of median doses followed similar patterns by diagnosis indicated on prescription (**Supplementary Figure 7**). For most medications, higher maximum doses were purchased less frequently than lower ones. No variation in the maximum dose was observed for fluoxetine (**Table 2**).

**Table 2.**
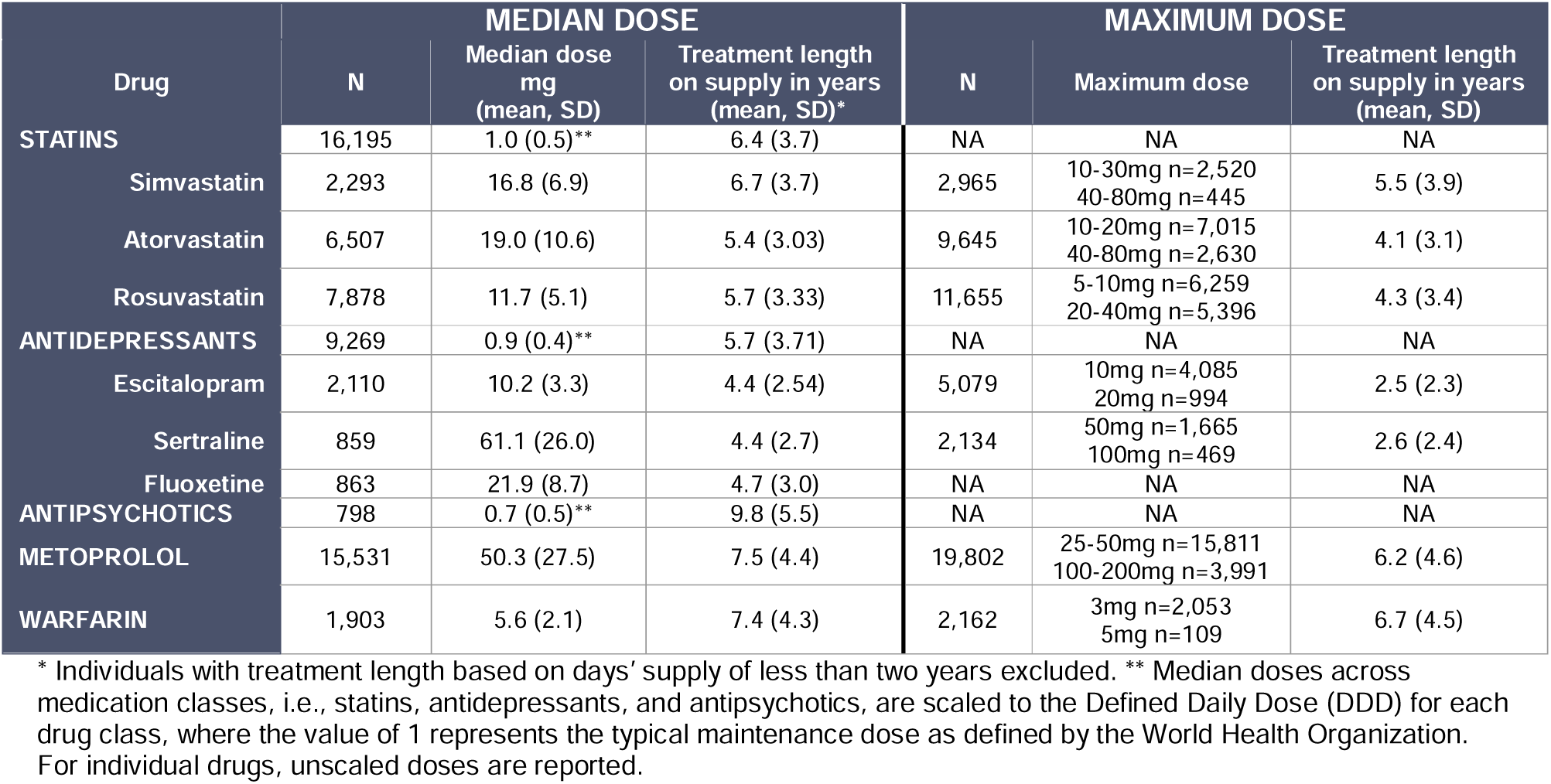
Overview of derived median and maximum dose variables.

The median and maximum dose variables partially recapitulated the associations observed with daily doses per purchase, though some differences emerged (**Figure 2B-C)**. CHD PGS and BMI PGS were similarly independently linked to higher median doses of statins, both at the class level (CHD PGS: β=0.02, SE=0.004, P=7.0×10^−10^; BMI PGS: β=0.01, SE=0.004, P=1.2×10^−4^) and for individual drugs, i.e., simvastatin (CHD PGS: β=0.03, SE=0.008, P=1.6×10^−4^; BMI PGS: β =0.03, SE=0.008, P=1.9×10^−4^), and rosuvastatin (CHD PGS: β=0.03, SE=0.005, P=7.3×10^−9^; BMI PGS: β=0.02, SE=0.005, P=0.001). For atorvastatin, only CHD PGS remained significant (**Supplementary Table 6**). Similarly, CHD PGS was solely associated with the maximum dose of individual statins (**Supplementary Table 7**). For metoprolol, BMI PGS and SBP PGS were linked to both median (BMI PGS: β=0.04, SE=0.004, P=1.8×10^−25^; SBP PGS: β =0.02, SE=0.004, P=1.3×10^−8^) and maximum doses (BMI PGS: OR=1.16, 95%CI 1.12–1.20, P=5.1×10^−15^; SBP PGS: OR=1.12, 95%CI 1.08–1.17, P=1.8×10^−9^), while BMI PGS correlated only with the median dose of warfarin (**Supplementary Table 6**). Among antidepressants, only fluoxetine showed a significant association with SBP PGS for its median dose (**Figure 2C**). Consistent with daily doses, EA PGS was inversely linked to the maximum doses of atorvastatin and rosuvastatin in individual models. Interestingly, MDD PGS, the primary indication for sertraline, showed a nominally significant positive association with its median dose, with an even more significant effect on maximum dose (**Supplementary Table 6-7**).

### Genome-wide association studies with medication dose variables

We next sought to identify genetic variants associated with the derived median and maximum dose variables across the genome. Two genome-wide scans were performed: the first adjusted for sex, birth year, treatment length on supply, and genotype PCs, while the second additionally included the relevant PGS for each drug (CHD for statins, MDD for antidepressants, SCZ for antipsychotics, SBP for metoprolol, and VTE for warfarin). The analyses revealed robust associations with metoprolol and warfarin in gene regions previously implicated in their metabolism. For metoprolol median dose, the strongest signal was observed in *CYP2D6* (rs5751229 A/G, 18 kb upstream from the gene, β=–0.11, P=1.1×10^−20^; **Figure 3A; Supplementary Figure 10A**) with consistent results with SBP PGS adjustment (**Supplementary Figure 8C**). This association was also detected for the maximum dose variable but with a weaker effect (OR=0.83, P=2.8×10^−9^; **Figure 3B**; **Supplementary** Figures 8D and 10B). Interestingly, when accounting for the loss-of-function SNV known to tag *CYP2D6*4* (rs3892097)^68^ in the metoprolol median dose GWAS, the association at *CYP2D6* was no longer genome-wide significant (rs5751229 A/G, β=–0.11, P=3.3×10JJ; **Supplementary Figure 8E**), indicating that the main signal was tagging the poor metabolizer genotype that results in a non-functional CYP2D6 enzyme. For warfarin median dose, top associations were identified within *VKORC1* (rs9934438 A/G, β=–0.69, P=4.2×10^−148^) and within *CYP2C9* (rs9332238 A/G, β =–0.53, P=8.9×10^−60^) (**Figure 3C; Supplementary Figure 10CD)**, with similar signals with VTE PGS adjustment (**Supplementary Figure 9C**). However, these associations were not observed for maximum dose (*VKORC1* rs9934438 A/G, OR=0.57, P=2.5×10^−5^; *CYP2C9* rs9332238 A/G, OR=0.58, P=0.003, in a model without VTE PGS; **Figure 3D**; **Supplementary** Figures 9D).

**Figure 3.**
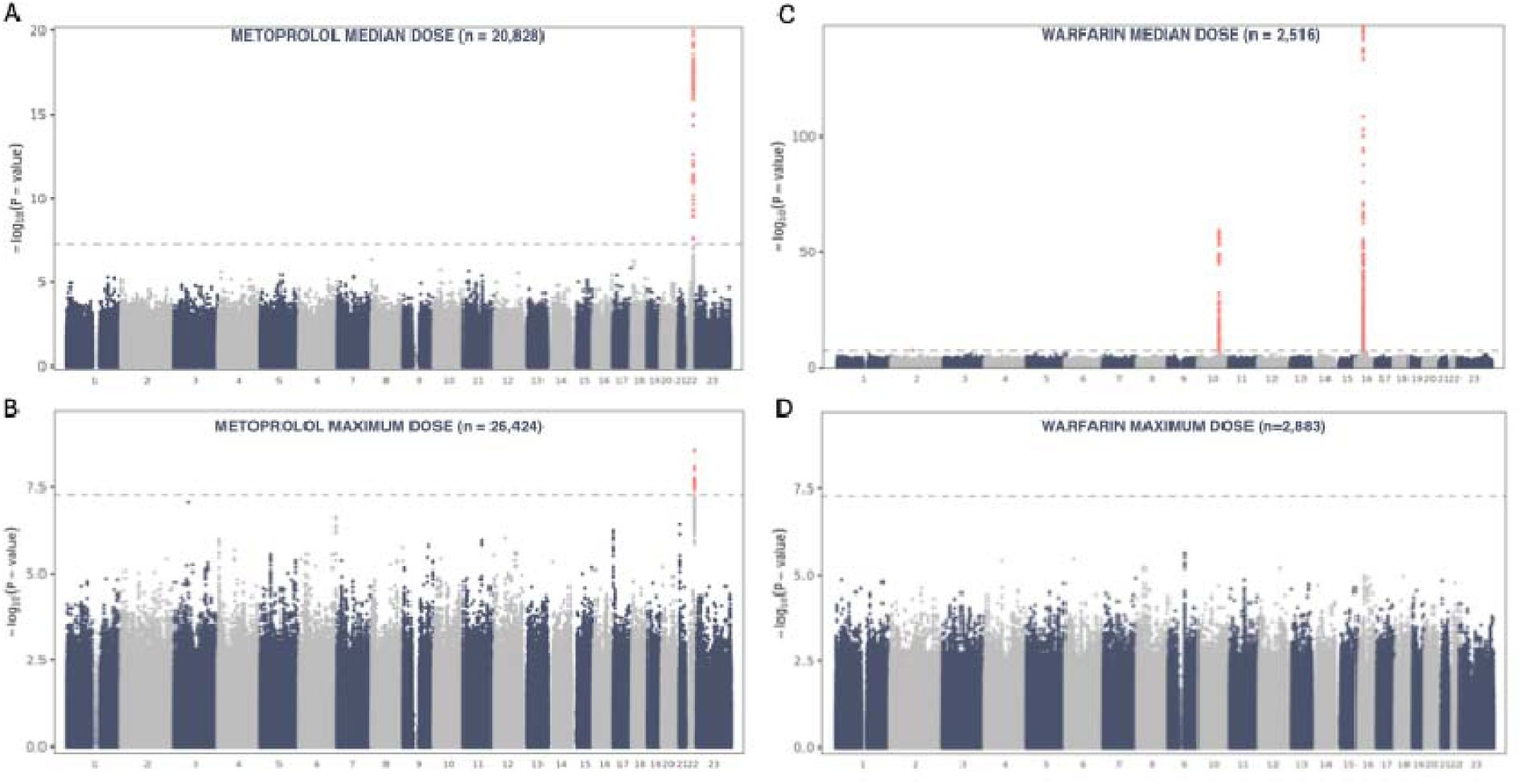
GWAS results for metoprolol and warfarin. Manhattan plot for metoprolol (A) median dose and (B) maximum dose adjusted for birth year, sex, treatment length on supply, and the first ten PCs. Manhattan plot for warfarin (C) median dose and (D) maximum dose adjusted for birth year, sex, treatment length on supply, and the first ten PCs. The y-axis represents −log_10_(P-values) for the association of SNVs with the dose variables, the horizontal dashed line indicates the genome-wide significance threshold (P<5×10^−8^), red dots genome-wide significant SNVs, and number 23 on x-axis denotes chromosome X.

No genome-wide significant associations were observed for other dose variables and medications (**Supplementary Note**, **Supplementary** Figures 11-19). While the top signal for atorvastatin maximum dose in the *LPA* locus on chromosome 6 (rs562851244 T/G, 1 kb upstream from the gene, OR=2.1, P=2.3×10^−7^; **Supplementary Figure 13C**) has previously been linked to statin use^17^ (EstBB: OR=1.1, P=2.6×10^−19^; FinnGen: OR=1.8, P=5.47×10^−15^) and coronary atherosclerosis in the pooled biobank GWAS (mvp-ukbb.finngen.fi: β=0.43, P=2.0×10^−15^), the association was substantially attenuated when accounting for CHD PGS (OR=1.4, P=0.02; **Supplementary Figure 13D**).

Given the absence of strong PGx signals for other medications, we assessed whether medication-specific PGx genes collectively showed enrichment of signal compared to what would be expected by chance. While PGx genes displayed suggestive evidence of associations in the GWAS of median statin dose at the class level without CHD PGS adjustment (P=0.08, adjusted P=0.11), distinct patterns emerged across individual statins. Significant drug-specific PGx signals for median dose were observed for simvastatin (P=0.01) and rosuvastatin (P=0.04) in the unadjusted GWAS only, whereas strong effects for maximum dose were detected for simvastatin and atorvastatin both without (P=0.045, P=0.03, respectively) and with CHD PGS adjustment (P=0.02, P=0.03, respectively). This indicates that PGx enrichment and its combined effects beyond CHD liability may be better detected at peak dosing while being diluted at lower levels captured by the median dose metric. No enrichment was found for antipsychotics, antidepressants or sertraline (**Supplementary Figure 20**).

The most prominent LD-pruned PGx genes surpassing empirical significance threshold of suggestive signal were *ABCB1* for simvastatin, and antipsychotics; *SLCO1B1* for statins and atorvastatin, reinforcing the well-established role of *SLCO1B1* in statin drug response; *SLCO2B1* for simvastatin, atorvastatin, and rosuvastatin, and *UGT2B7* for rosuvastatin and sertraline. While *CYP4F2* ranked among the top PGx genes associated with the median dose for warfarin, its effect was considerably weaker compared to the primary PGx signals (*VKORC1* and *CYP2C9)*, confirming its secondary yet relevant role in warfarin dose response. For warfarin maximum dose, *CYP3A4* ranked among top PGx genes with *VKORC1* and *CYP2C9* (**Supplementary Table 8, Supplementary Figure 21**). Overall, these findings are consistent with established pharmacogenetic knowledge, validating known drug-gene interactions and indicating that the method used to define drug dose phenotypes effectively captures relevant PGx signals. Top five variants in GWAS results for each PGx gene without LD pruning are outlined in **Supplementary Table 9**.

## DISCUSSION

While real-world health data has opened new avenues in human genomics, their application in PGx has remained modest. In this study, we explored the feasibility of using drug purchase data to derive medication doses and evaluate their variability in relation to genetic factors. Through PGS analyses, GWAS, and PGx gene enrichment testing, we examined a diverse range of cardiovascular and psychiatric medications using EHR data from EstBB. Our findings highlighted that while drug purchase data is suitable for this purpose, as evidenced by strong expected associations for metoprolol and warfarin, genetic associations partially reflect the nature of EHR data, capturing both biological and healthcare-related influences. The detection of strong PGx signals beyond underlying trait predisposition underscores the potential of real-world health data to deepen our understanding of treatment response.

The analyses of polygenic liabilities for complex traits revealed distinct patterns in genetic associations. First, robust genetic signals for indicator traits, such as CHD PGS for statin dosing and SBP PGS for metoprolol dosing, indicate that individuals with a higher genetic predisposition to these conditions are more likely to receive higher doses, reflecting both disease liability and clinical treatment strategies. Interestingly, CHD PGS and BMI PGS showed independent associations with medication doses, even after accounting for treatment changes following cardiovascular events. This underscores that these pooled genetic liabilities capture complementary molecular mechanisms contributing to higher dose requirements. Second, the association of BMI PGS with multiple medications suggests that prescribing decisions may be influenced by BMI as a health indicator. Similarly, the inverse association between SBP PGS and fluoxetine dose, and between HDL PGS and metoprolol dose, may reflect clinicians’ treatment preference based on cardiovascular risk considerations. Third, the associations between EA PGS with statins and antidepressants could be attributed to patient behaviour. For statins, the negative association may reflect broader genetic influences such as higher proactivity in managing their health, healthier lifestyle choices, and access to healthcare resources (e.g., employer-supported healthcare visits). On the other hand, the positive association with antidepressant dose could suggest better self-care behaviours and a greater likelihood of seeking medical treatment for mental health conditions.

Genome-wide scans revealed strong PGx signals for metoprolol and warfarin, as well as suggestive evidence for statins both at the class level and the individual drug level. These signals persisted even after accounting for underlying trait genetics, highlighting the independent contribution of PGx to medication dosing and that genetic determinants of drug metabolism and transport are not fully captured by the PGSs of the indicator trait. This underscores the limited utility of relying solely on trait PGS for predicting drug response.

Furthermore, while cardiovascular drugs have clinical tests for monitoring treatment effect, for instance prothrombin time for warfarin, cholesterol reduction for statins, and blood pressure for metoprolol, such dose optimization usually takes weeks to months. For antidepressants or antipsychotics, therapeutic drug monitoring is more commonly used to determine a patient’s drug exposure at standard starting doses, thereby facilitating more rapid dose optimization. However, such tests require time and resources of healthcare systems, reinforcing the value of pre-emptive PGx testing. While for warfarin, dose calculators have been developed that integrate clinical information (including drug-drug interactions) and genetic information for *CYP2C9* and *VKORC1* and in some models, for *CYP4F2*^69^, such models are not routinely applied since genetic data is rarely available in routine medical care. Well-powered GWAS of drug dosing could enhance these tools even further by integrating a broader dose score index that pools the effects across multiple genes, similar to PGS. Given the increasing clinical content and low cost of genotyping microarrays, such pre-emptive genetic tests can enable both PGS-based risk estimation as well as PGx reports, facilitating more precise and personalized treatment strategies.

The necessity of accounting for underlying genetics in the GWAS of medication dose was further illustrated for atorvastatin, where a significant *LPA* signal disappeared after adjusting for CHD PGS. While the *LPA* locus has been robustly associated with higher CHD risk^57,70^, likely explaining the attenuation of the signal, it has also been shown to modulate LDL response to statins^24,71^. As standard LDL-C assays do not distinguish between cholesterol in LDL and lipoprotein(a) particles, these observed associations could primarily reflect differences in lipoprotein(a) levels rather than a direct effect on statin metabolism, as statins do not influence lipoprotein(a) concentrations^72^. For other medications, adjusting for the polygenic liability of the indicator diagnosis did not attenuate GWAS signals, likely due to modest associations between the genetics of the underlying trait and dosing variability. The lack of enrichment for antipsychotics and antidepressants could reflect diluted PGx effects when analysed at the drug class level as well as highlight the need for larger sample sizes to identify dose-affecting PGx variants.

EHR data presents several limitations that impact the derivation of medication doses. Daily doses per purchase can be affected by factors such as stocking up or long breaks in treatment, while median doses as a consolidated metric likely underestimate typical maintenance doses. Similarly, the length of treatment use periods and different filtering criteria can affect dose derivation. Interestingly, different strategies revealed different associations. For instance, GWAS identified strong associations for metoprolol and warfarin with median dose, while PGx effects for statins were better detectible when testing maximum dose. Similarly, both metrics showed an association with MDD PGS for sertraline, whereas linear mixed modelling with daily doses did not detect this relationship, emphasizing the importance of tailored approaches in capturing medication doses. Additionally, the use of Defined Daily Doses to scale medications at the class level may not be optimal for capturing drug-specific dose variability. Furthermore, the minimal polygenic signals observed in GWAS align with previous studies reporting low heritability for drug response^5,19,24,25^, and that large-effect variants likely contribute little to the heritability of PGx phenotypes^73^. The limited sample sizes for individual drugs further emphasize the need to improve statistical power for discovery as well as for detecting enrichment across all known PGx genes.

In conclusion, this study is among the first to derive medication doses from biobank-linked medication purchase data to explore both polygenic and known PGx associations at scale. The thorough analysis of derived drug dose estimates underscores the importance of systematic efforts to account for the complexities of real-word data, and highlights that associated genetic signals can reflect not only disease predispositions and pharmacokinetic effects but also broader clinical prescribing practices and patients’ healthcare-seeking behaviours. Our findings collectively highlight the need to integrate pharmacogenomic and polygenic insights to refine dose optimization and advance precision medicine. Moving forward, the possibility of combining all relevant factors with pre-emptive genetic testing offers new opportunities for more equitable personalized therapy.

## Data Availability

GWAS summary statistics are publicly available as provided in respective publications. Individual-level data at EstBB can only be accessed through EstBB. Further info about access is provided at https://genomics.ut.ee/en/content/estonian-biobank.

## CONTRIBUTORS

Conceptualisation: L.M., M.A., L.B.L., S.K. Data curation: M.A., S.K., L.B.L., K.K., E.B.R.T. Formal analysis: L.B.L., S.K., M.A. Funding acquisition: L.M., M.A. Investigation: L.B.L., S.K., K.K., M.A., L.M. Methodology: L.B.L., S.K., M.M., M.A. Supervision: M.A., L.M. Visualisation: L.B.L., S.K., K.K., M.A. Writing: original draft: M.A., L.M., L.B.L., S.K., K.K. Writing—review & editing: M.A., L.M., L.B.L., S.K., K.K., M.M.

## DECLARATION OF INTERESTS

All authors declare no competing interests.

## ACKNOWLEDGEMENTS

This research was supported by EU Horizon 2020 Research and Innovation Programme under Grant agreement No. 964874 (Realment), by the Estonian Research Council’s grant PSG1028, grant PRG2625, grant PRG1197, and grant PRG1911, by the Estonian Ministry of Education and Research’s grant TK214, by the Estonian Center of Genomics/Roadmap II grant TT17 funded by the Estonian Research Council, by the Swedish Research Council’s grant No. 2021-02732, and by the University of Tartu grant PLTGI24925. The authors acknowledge the Estonian Biobank Research Team (Andres Metspalu, Tõnu Esko, Reedik Mägi, Mait Metspalu, Mari Nelis, Georgi Hudjashov), and Urmo Võsa for the parallelized PGS calculation pipeline. Data analysis was carried out in part in the High-Performance Computing Center of University of Tartu.

## Notes

### Competing Interest Statement

The authors have declared no competing interest.

### Author Declarations

The study was approved by the Estonian Committee on Bioethics and Human Research at the Estonian Ministry of Social Affairs (24 March 2020, nr 1.1-12/624).

