## Supplementary Figures for "Investigating the genetic factors of medication dosing using biobank-linked drug purchase data": Supplementary_Figures_dose_genetics_submit.pdf

### SUPPLEMENTARY NOTE

Despite no genome-wide significant associations for the median and maximum dose of statins, simvastatin, atorvastatin, rosuvastatin, antidepressants, escitalopram, sertraline, fluoxetine, and antipsychotics, several loci showed suggestive signals.

For simvastatin median dose, a signal was observed on chromosome 18 (rs16956046 A/C, $\beta$=–0.20, P=$2.1\times10^{-7}$) near *VAPA*, involved in lipid transport and cellular cholesterol distribution[1,2] (**Supplementary Figure 12A**). For atorvastatin maximum dose, in addition to the *LPA* locus on chromosome 6, a suggestive association lies on chromosome X near *SMARCA1* (rs4830094 C/A, OR=1.15, P=$7.9\times10^{-7}$; **Supplementary Figure 13C**), linked with essential hypertension in pooled biobank GWAS (mvp-ukbb.finngen.fi: $\beta$=–0.01, P=$5.9\times10^{-4}$). For rosuvastatin median dose, signals were detected on chromosome 21 near *RUNX1* (rs2834862 C/G, $\beta$=–0.09, P=$6.4\times10^{-8}$), on chromosome 1 within *DISP1* (rs1890619 A/G, $\beta$=0.17, P=$2.3\times10^{-7}$), which is linked with ischemic heart disease in pooled biobank GWAS (mvp-ukbb.finngen.fi: $\beta$=–0.02, P=$1.5\times10^{-5}$), and on chromosome 12 within *CEP83* (rs2176899 T/A, $\beta$=–0.07, P=$6.7\times10^{-7}$), previously associated with type 2 diabetes (mvp-ukbb.finngen.fi: $\beta$=0.01, P=$2.6\times10^{-6}$) (**Supplementary Figure 14A**).

For antidepressant median dose, a suggestive locus was identified on chromosome 4 (rs111809139 T/G, $\beta$=–0.22, P=$4.8\times10^{-7}$) within *LINC02267* (**Supplementary Figure 15A**). For escitalopram median dose, an association was found on chromosome 3 near *ETV5* (rs13095986 A/G, $\beta$=0.15, P=$1.5\times10^{-7}$; **Supplementary Figure 16A**), linked to obesity (mvp-ukbb.finngen.fi: $\beta$=0.02, P=$1.0\times10^{-8}$), and for escitalopram maximum dose on chromosome 15 within *MYO1E* (rs3794491 G/A, OR=1.31, P=$2.4\times10^{-7}$; **Supplementary Figure 16C**), annotated to a CpG site shown to be associated with self-reported antidepressant use[3]. For sertraline maximum dose, suggestive peaks were found on chromosome 1 within *PTPN14* (rs6700380 T/C, OR=0.66, P=$7.4\times10^{-8}$) and in chromosome 10 within *PLXDC2* (rs9651366 G/A, OR=1.77, P=$4.2\times10^{-7}$), residing within a locus previously associated with response to antidepressant treatment[4] (**Supplementary Figure 17C**). For antipsychotic median dose, a locus was identified on chromosome 4 near *SCLT1* (rs4263417, T/C, $\beta$=–0.18, P=$2.3\times10^{-7}$; **Supplementary Figure 19A**).

For metoprolol maximum dose, suggestive associations were observed on chromosome 6 within *RPS6KA2* (rs9459731 C/G, OR=0.70, P=$2.5\times10^{-7}$), and on chromosome 17 within *RPH3AL* (rs7222726 A/C, $\beta$=0.83, P=$5.7\times10^{-7}$; **Figure 3B**). For warfarin maximum dose, a sub-threshold peak was observed on chromosome 9 within *TRPM3* (rs7875827 C/T, OR=1.75, P=$2.3\times10^{-6}$; **Figure 3D**), linked with type 2 diabetes in pooled biobank GWAS (mvp-ukbb.finngen.fi: $\beta$=0.01, P=$5.6\times10^{-6}$). For warfarin median dose, an SNV on chromosome 2 (rs140698681 CTTTTG/C, $\beta$=-0.61, P=$2.66\times10^{-8}$, MAF 0.01) within *IL1RL2* surpassed the GWAS significance threshold, but lacked LD-supported signals, suggesting caution in interpretation (**Figure 3C)**.

### SUPPLEMENTARY FIGURES

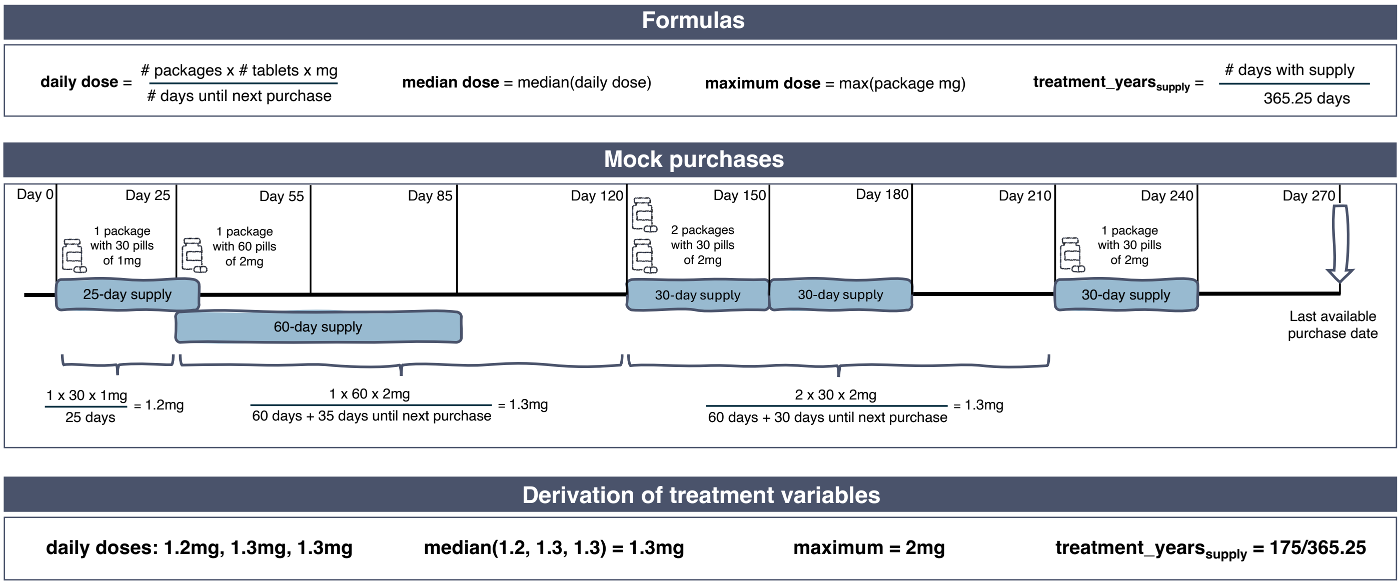

**Supplementary Figure 1. Overview of the derivation of treatment variables from drug purchase data.** The top panel provides the formulas used for derivation, the middle panel depicts a mock example of purchase data, and the lower panel shows the derived treatment variables based on the mock example. The middle panel displays four purchases for an individual over a 270-day period: 1 package of 30 tablets of 1mg, 1 package of 60 tablets of 1mg, 2 packages of 30 tablets of 2mg, and 1 package of 30 tablets of 1mg. Below each purchase, the lower sections indicate the corresponding values used for the derivation of the daily dose. Of note, the second purchase of 1 package of 60 tablets of 2mg is depicted to occur 5 days before the end of the supply of the first purchase. Specifically, the daily dose per purchase was calculated by multiplying the package content (dose in mg and number of pills) by the number of packages bought and dividing by the number of days until the next purchase. The median dose variable was calculated by taking the median of all derived daily doses. The maximum dose variable was defined as the highest purchased dose (mg per package), requiring at least three purchases of the same dose to capture sustained higher-dose use. For analyses, binary variables for maximum dose contrasted the highest dose in milligrams with all other doses.

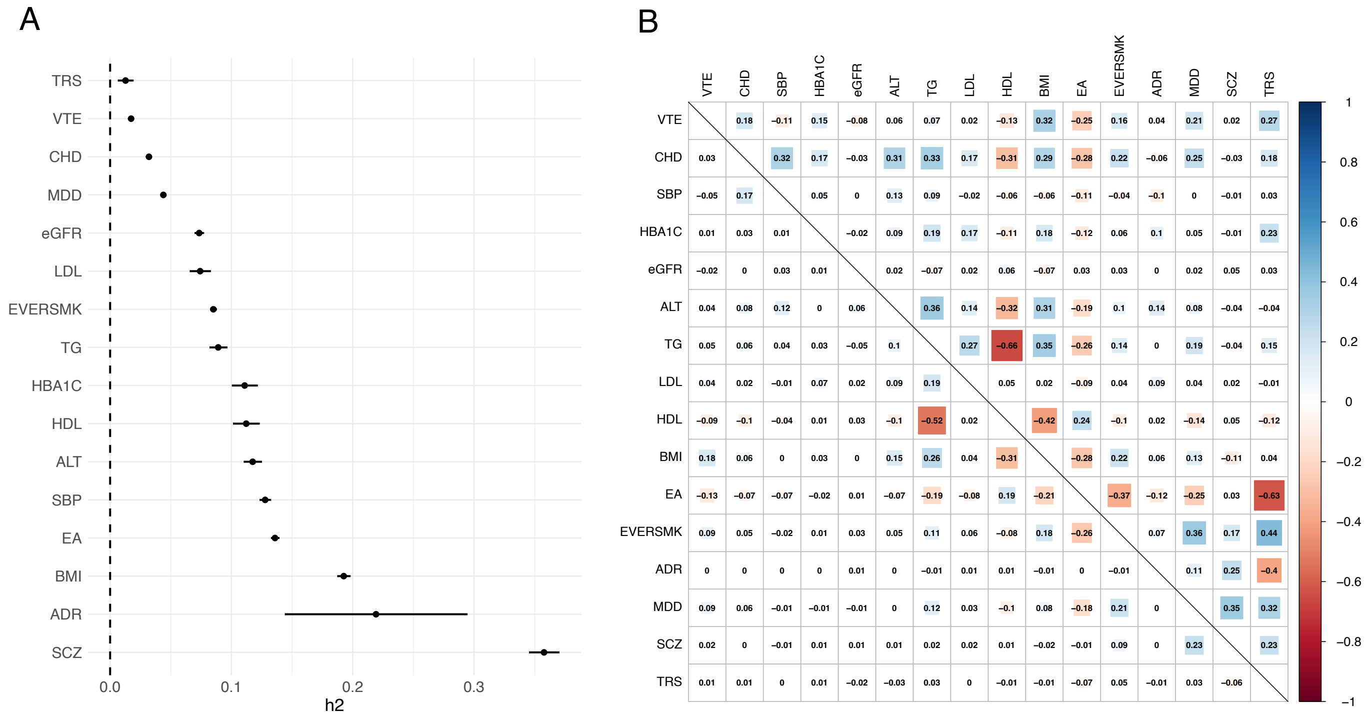

**Supplementary Figure 2. Heritability estimates and correlations of traits considered for PGS analysis.** (A) The observed heritability estimates were calculated using LDSC based on the respective GWAS summary statistics. (B) Correlation matrix of the underlying genetics of the traits used for PGS analysis. The upper triangle outlines the genetic correlations retrieved with LDSC using published GWAS summary statistics. The lower triangle indicates the Pearson correlation estimates of the PGSs calculated using all unrelated (PLINK PI_HAT <0.2) EstBB genotype individuals of European ancestry (n=114,346). While the correlation estimates between the two approaches align, the correlations among PRSs are notably lower.

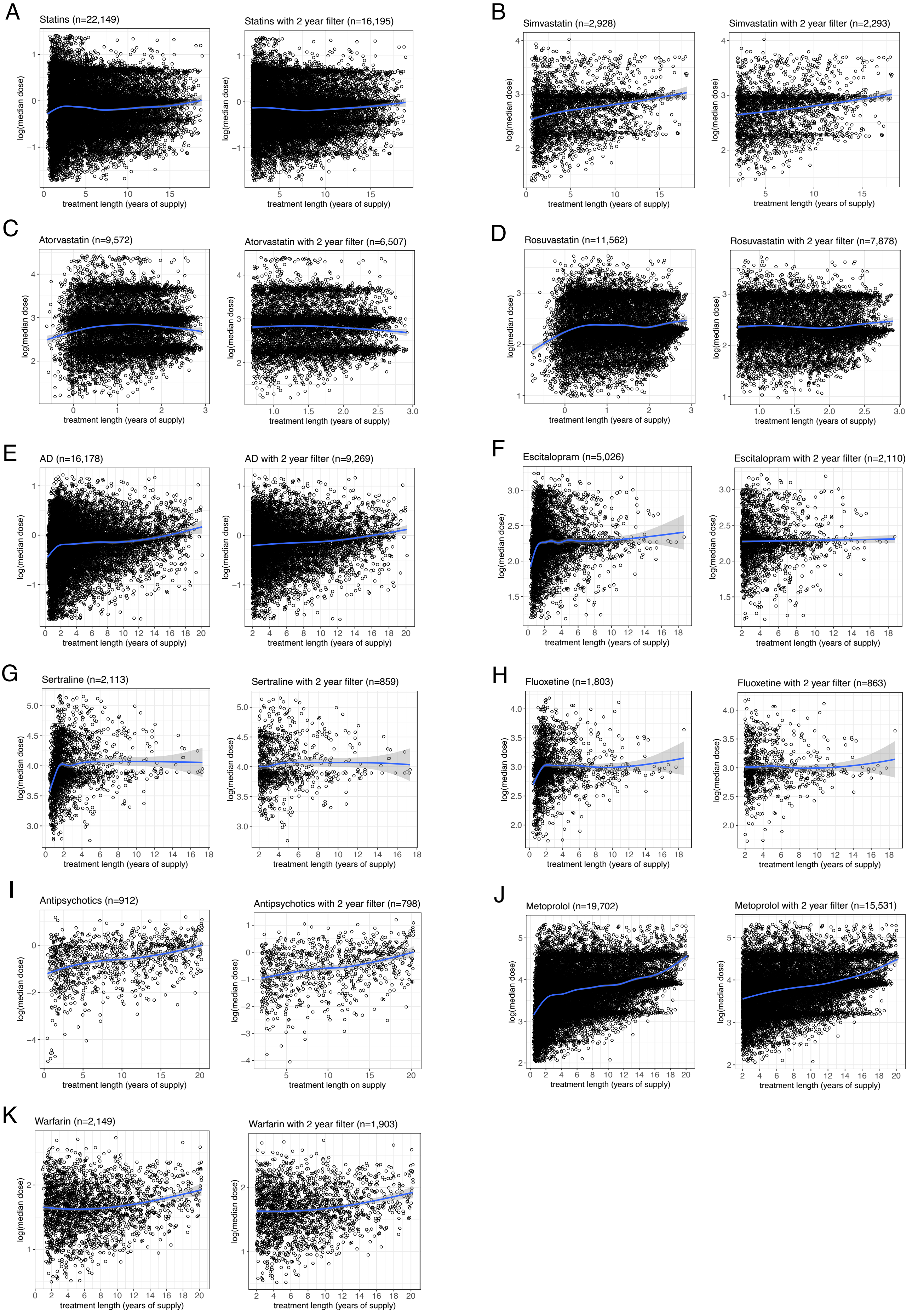

**Supplementary Figure 3. Association between the derived median doses across purchases and treatment length based on supply.** Associations are shown for all derived median doses and for those filtered to include at least 2 years of treatment based on supply, stratified by (A) statins, (B) simvastatin, (C) atorvastatin, (D) rosuvastatin, (E) antidepressants (AD, subset with F32, F33, F41.2), (F) escitalopram, (G) sertraline, (H) fluoxetine, (I) antipsychotics (subset with F20-F29), (J) metoprolol, and (K) warfarin. The blue fitted line represents a LOESS-smoothed trend illustrating the relationship between treatment length and median dose.

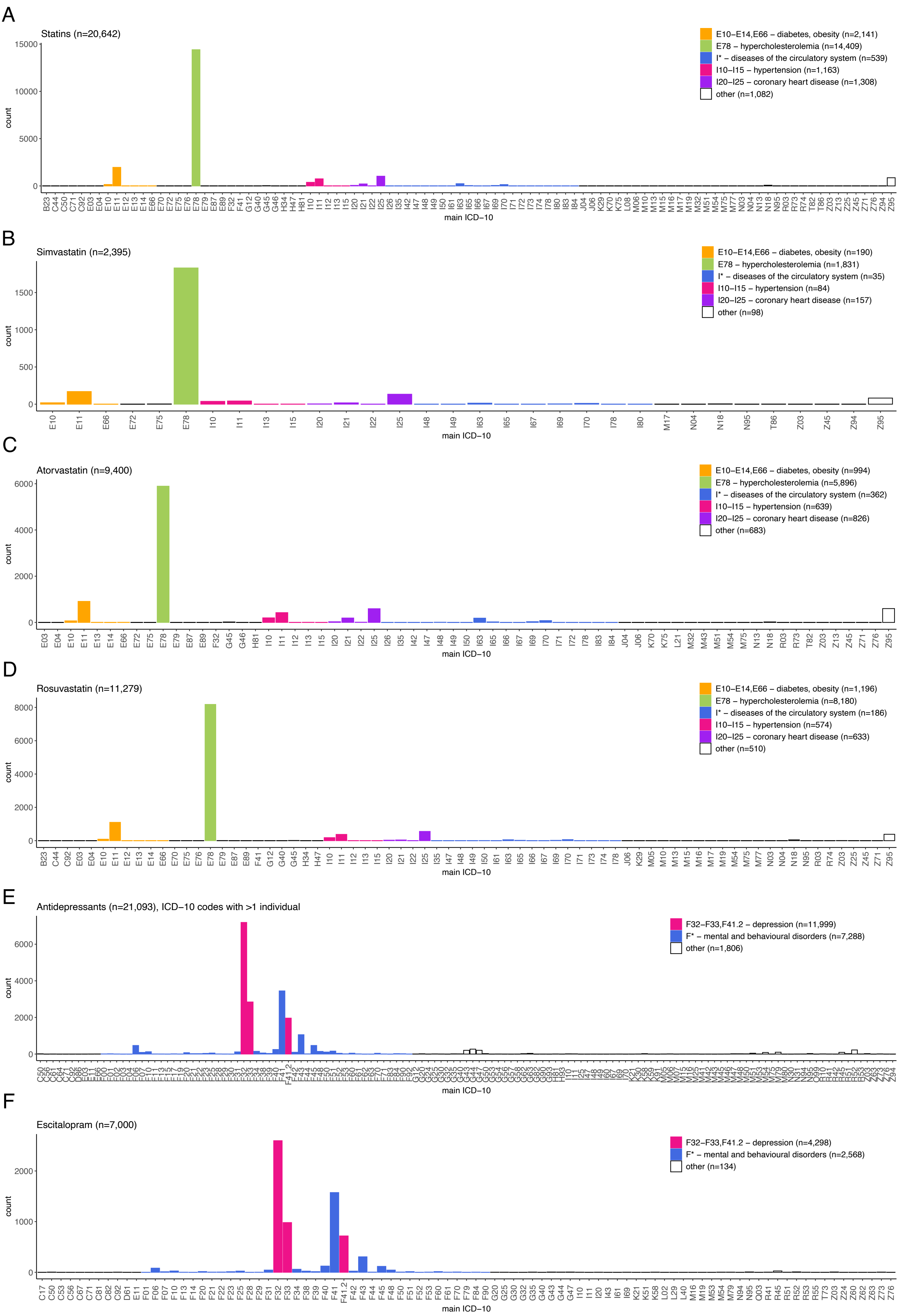
A
Statins (n=20,642)
E10–E14,E66 – diabetes, obesity (n=2,141)
E78 – hypercholesterolemia (n=14,409)
I* – diseases of the circulatory system (n=539)
I10–I15 – hypertension (n=1,163)
I20–I25 – coronary heart disease (n=1,308)
other (n=1,082)
count
main ICD−10
B
Simvastatin (n=2,395)
E10–E14,E66 – diabetes, obesity (n=190)
E78 – hypercholesterolemia (n=1,831)
I* – diseases of the circulatory system (n=35)
I10–I15 – hypertension (n=84)
I20–I25 – coronary heart disease (n=157)
other (n=98)
count
main ICD−10
C
Atorvastatin (n=9,400)
E10–E14,E66 – diabetes, obesity (n=994)
E78 – hypercholesterolemia (n=5,896)
I* – diseases of the circulatory system (n=362)
I10–I15 – hypertension (n=639)
I20–I25 – coronary heart disease (n=826)
other (n=683)
count
main ICD−10
D
Rosuvastatin (n=11,279)
E10–E14,E66 – diabetes, obesity (n=1,196)
E78 – hypercholesterolemia (n=8,180)
I* – diseases of the circulatory system (n=186)
I10–I15 – hypertension (n=574)
I20–I25 – coronary heart disease (n=633)
other (n=510)
count
main ICD−10
E
Antidepressants (n=21,093), ICD−10 codes with >1 individual
F32–F33,F41.2 – depression (n=11,999)
F* – mental and behavioural disorders (n=7,288)
other (n=1,806)
count
main ICD−10
F
Escitalopram (n=7,000)
F32–F33,F41.2 – depression (n=4,298)
F* – mental and behavioural disorders (n=2,568)
other (n=134)
count
main ICD−10

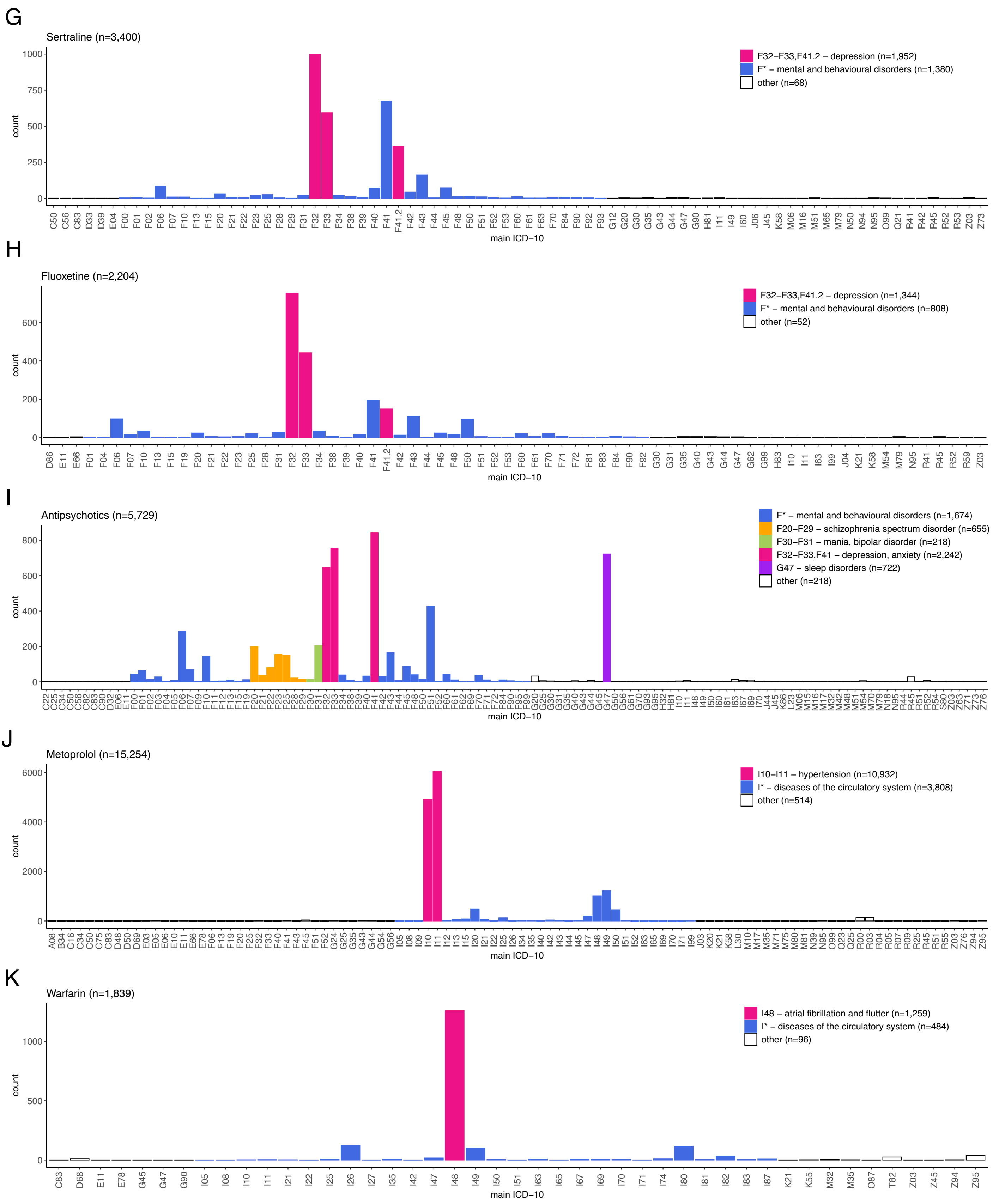

**Supplementary Figure 4. Number of individuals with drug purchases stratified by the primary ICD-10 code on prescription.** The counts are shown for (A) statins, (B) simvastatin, (C) atorvastatin, (D) rosuvastatin, (E) antidepressants (restricted to individuals with >1 purchase with the given ICD-10 code on prescription for figure readability), (F) escitalopram, (G) sertraline, (H) fluoxetine, (I) antipsychotics, (J) metoprolol, and (K) warfarin. ICD-10 codes are colour-coded by diagnostic groups to highlight the most prevalent ICD-10 codes and endpoints by drug. Of note, the sample sizes for individuals taking antidepressants, escitalopram, sertraline, and fluoxetine with F32, F33, F41.2 on prescription (E-H) and individuals taking antipsychotics with F20-F29 on prescriptions (I) in the figure differ from the sample set used in association testing (Table 1). This discrepancy stems from the 3SD filter (exclusion of doses deviating >3SDs from the log-scale mean), and the related sample exclusion (exclusion of one individual per related pair). These filters were applied across all drug users (in figure) or across subgroup (for association testing to maximize the number of cases).

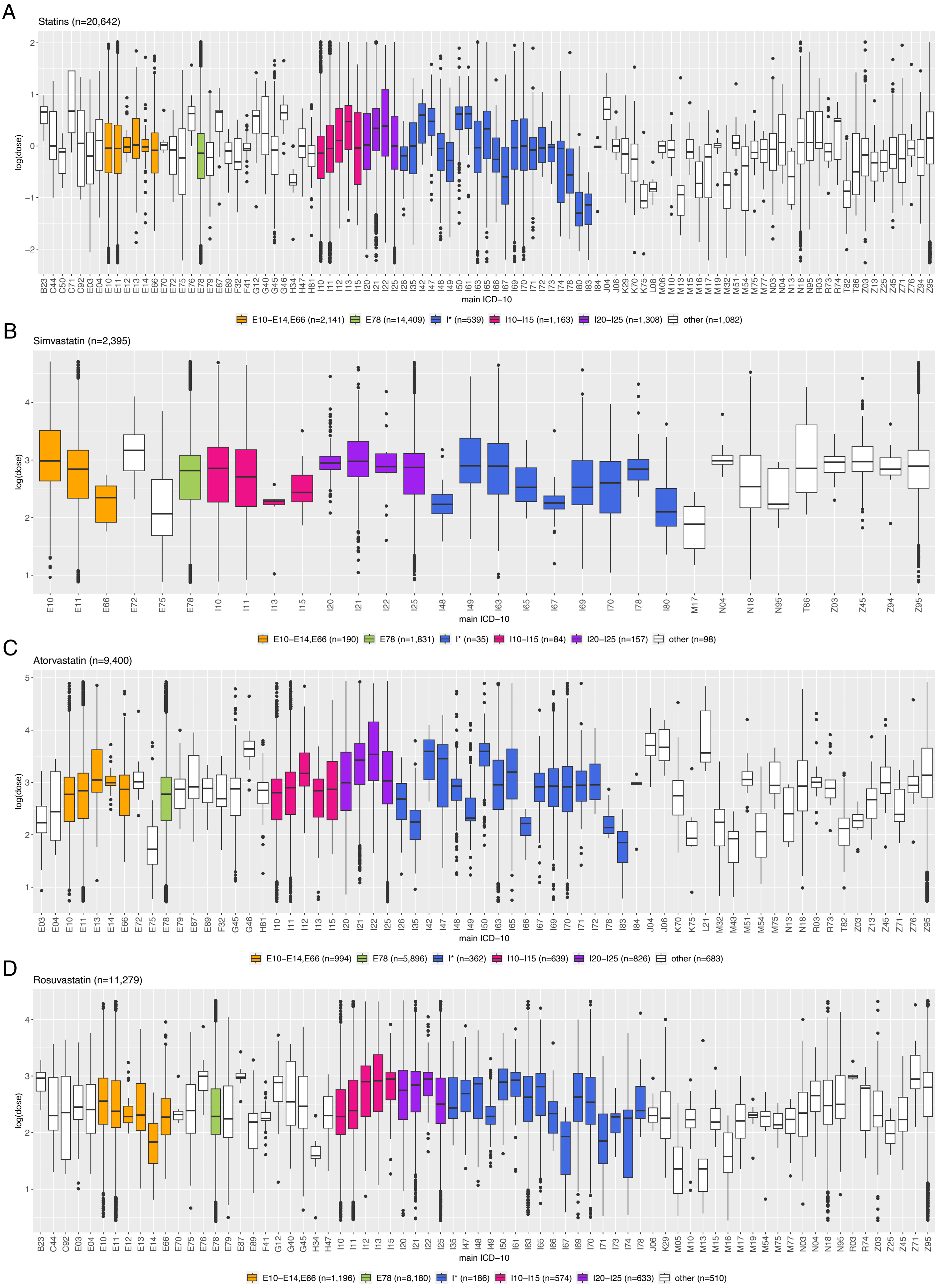
A
Statins (n=20,642)
log(dose)
main ICD−10
E10−E14,E66 (n=2,141)
E78 (n=14,409)
I* (n=539)
I10−I15 (n=1,163)
I20−I25 (n=1,308)
other (n=1,082)
B
Simvastatin (n=2,395)
log(dose)
main ICD−10
E10−E14,E66 (n=190)
E78 (n=1,831)
I* (n=35)
I10−I15 (n=84)
I20−I25 (n=157)
other (n=98)
C
Atorvastatin (n=9,400)
log(dose)
main ICD−10
E10−E14,E66 (n=994)
E78 (n=5,896)
I* (n=362)
I10−I15 (n=639)
I20−I25 (n=826)
other (n=683)
D
Rosuvastatin (n=11,279)
log(dose)
main ICD−10
E10−E14,E66 (n=1,196)
E78 (n=8,180)
I* (n=186)
I10−I15 (n=574)
I20−I25 (n=633)
other (n=510)

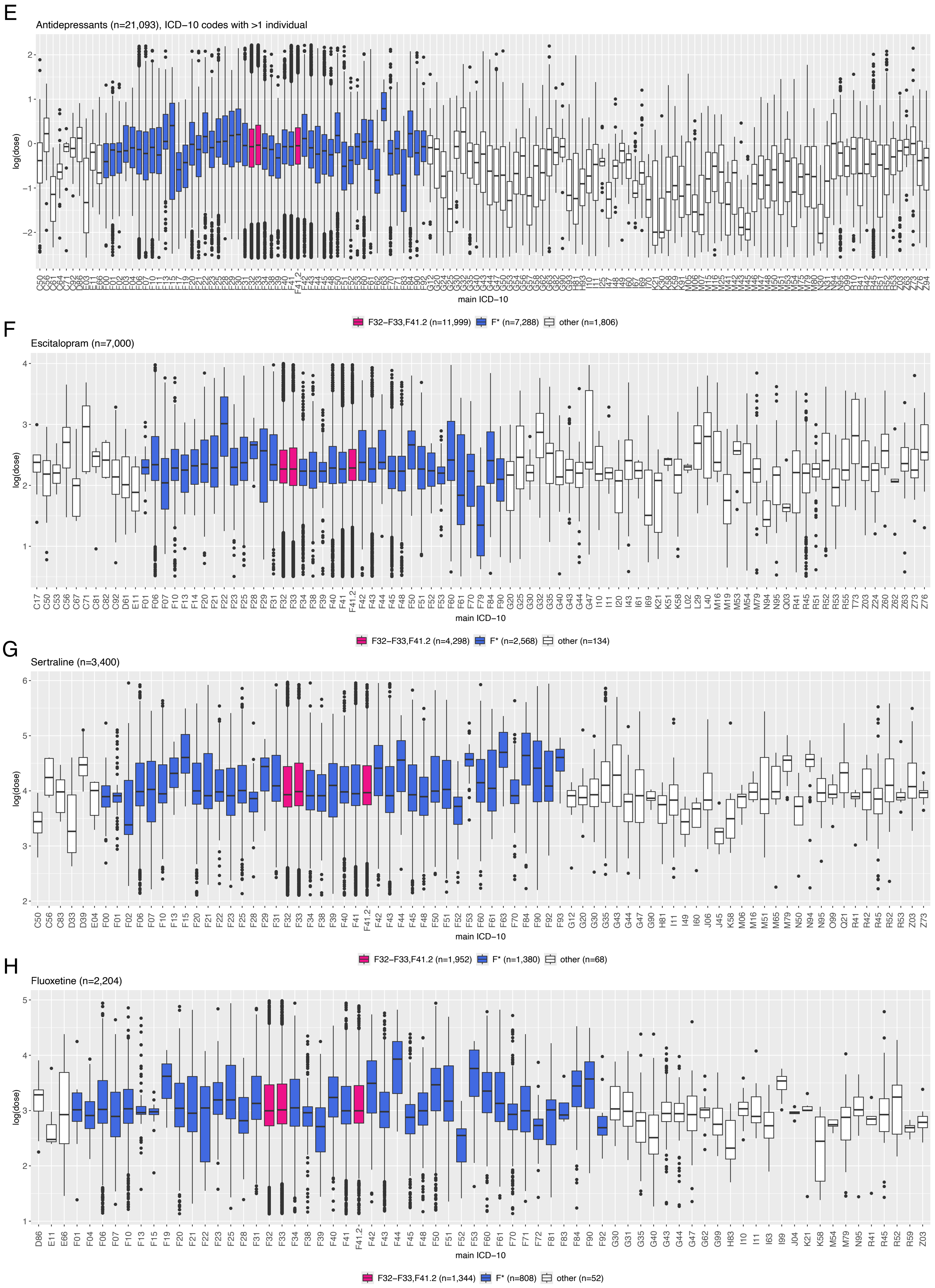
E
Antidepressants (n=21,093), ICD−10 codes with >1 individual
log(dose)
main ICD−10
F32−F33,F41.2 (n=11,999)
F* (n=7,288)
other (n=1,806)
F
Escitalopram (n=7,000)
log(dose)
main ICD−10
F32−F33,F41.2 (n=4,298)
F* (n=2,568)
other (n=134)
G
Sertraline (n=3,400)
log(dose)
main ICD−10
F32−F33,F41.2 (n=1,952)
F* (n=1,380)
other (n=68)
H
Fluoxetine (n=2,204)
log(dose)
main ICD−10
F32−F33,F41.2 (n=1,344)
F* (n=808)
other (n=52)

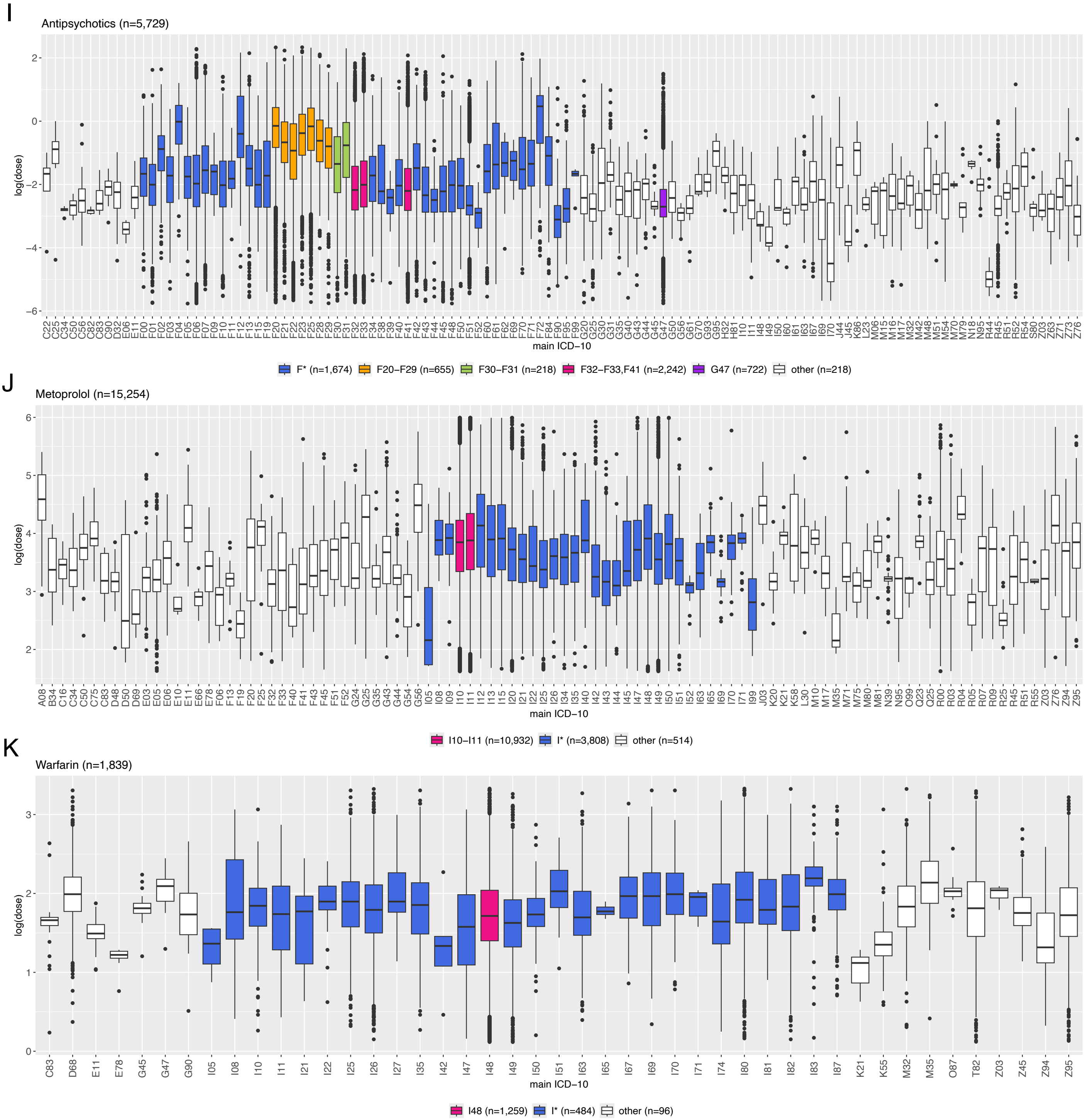

**Supplementary Figure 5. Overview of the distribution of derived daily doses per purchase on log scale by the primary ICD-10 code on prescription.** The distributions are outlined for (A) statins, (B) simvastatin, (C) atorvastatin, (D) rosuvastatin, (E) antidepressants (restricted to individuals with >1 purchase with the given ICD-10 code on prescription for figure readability), (F) escitalopram, (G) sertraline, (H) fluoxetine, (I) antipsychotics, (J) metoprolol, and (K) warfarin. ICD-10 codes are colour-coded by diagnostic groups to highlight the most prevalent ICD-10 codes and endpoints by drug. Specifically, for statins, simvastatin, atorvastatin, and rosuvastatin: E10-E14, E66 – diabetes, obesity; E78 – hypercholesterolemia; I10-I15 – hypertension; I20-I25 – coronary heart disease; I* – diseases of the circulatory system. For antidepressants, escitalopram, sertraline, and fluoxetine: F32-F33, F41.2 – depression; F* – mental and behavioural disorders. For antipsychotics: F20-F29 – schizophrenia spectrum disorder; F30-F31 – mania, bipolar disorder; F32-F33, F41 – depression, anxiety; G47 – sleep disorders; F* – mental and behavioural disorders. For metoprolol: I10-I11 – essential hypertension and hypertensive heart disease; I* – diseases of the circulatory system. For warfarin: I48 – atrial fibrillation and flutter; I* – diseases of the circulatory system. Of note, the sample sizes for individuals taking antidepressants, escitalopram, sertraline, and fluoxetine with F32, F33, F41.2 on prescription (E-H) and individuals taking antipsychotics with F20-F29 on prescriptions (I) in the figure differ from the sample set used in association testing (Table 1). This discrepancy stems from the 3SD filter (exclusion of doses deviating >3SDs from the log-scale mean), and the related sample exclusion (exclusion of one individual per related pair). These filters were applied across all drug users (in figure) or across subgroup (for association testing to maximize the number of cases).

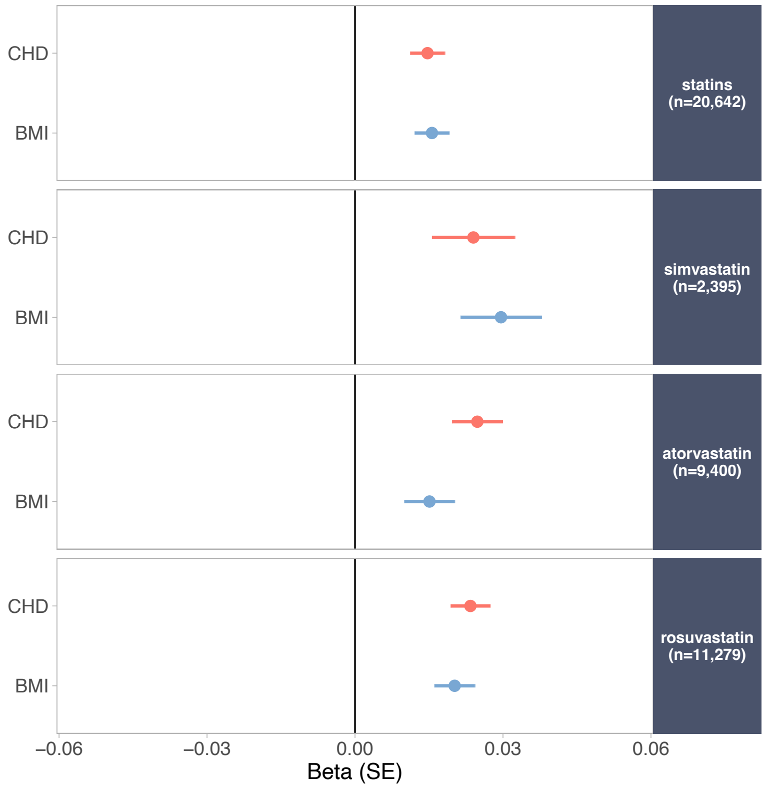

**Supplementary Figure 6. Effect sizes for PGSs significantly associated with derived doses for statins, simvastatin, atorvastatin, and rosuvastatin with disease diagnosis included in the model.** Only PGSs that surpassed Bonferroni correction and were independently associated are shown. Statins: CHD PGS $\beta$=0.02, SE=0.004, P=$2.2\times10^{-5}$; BMI PGS $\beta$=0.02, SE=0.004, P=$6.6\times10^{-6}$. Simvastatin: CHD PGS $\beta$=0.02, SE=0.008, P=$4.6\times10^{-3}$; BMI PGS $\beta$=0.03, SE=0.008, P=$3.3\times10^{-4}$. Atorvastatin: CHD PGS $\beta$=0.02, SE=0.005, P=$1.2\times10^{-6}$; BMI PGS $\beta$=0.02, SE=0.005, P=$2.8\times10^{-3}$. Rosuvastatin: CHD PGS $\beta$=0.02, SE=0.004, P=$4.6\times10^{-9}$; BMI PGS $\beta$=0.02, SE=0.004, P=$7.6\times10^{-7}$.

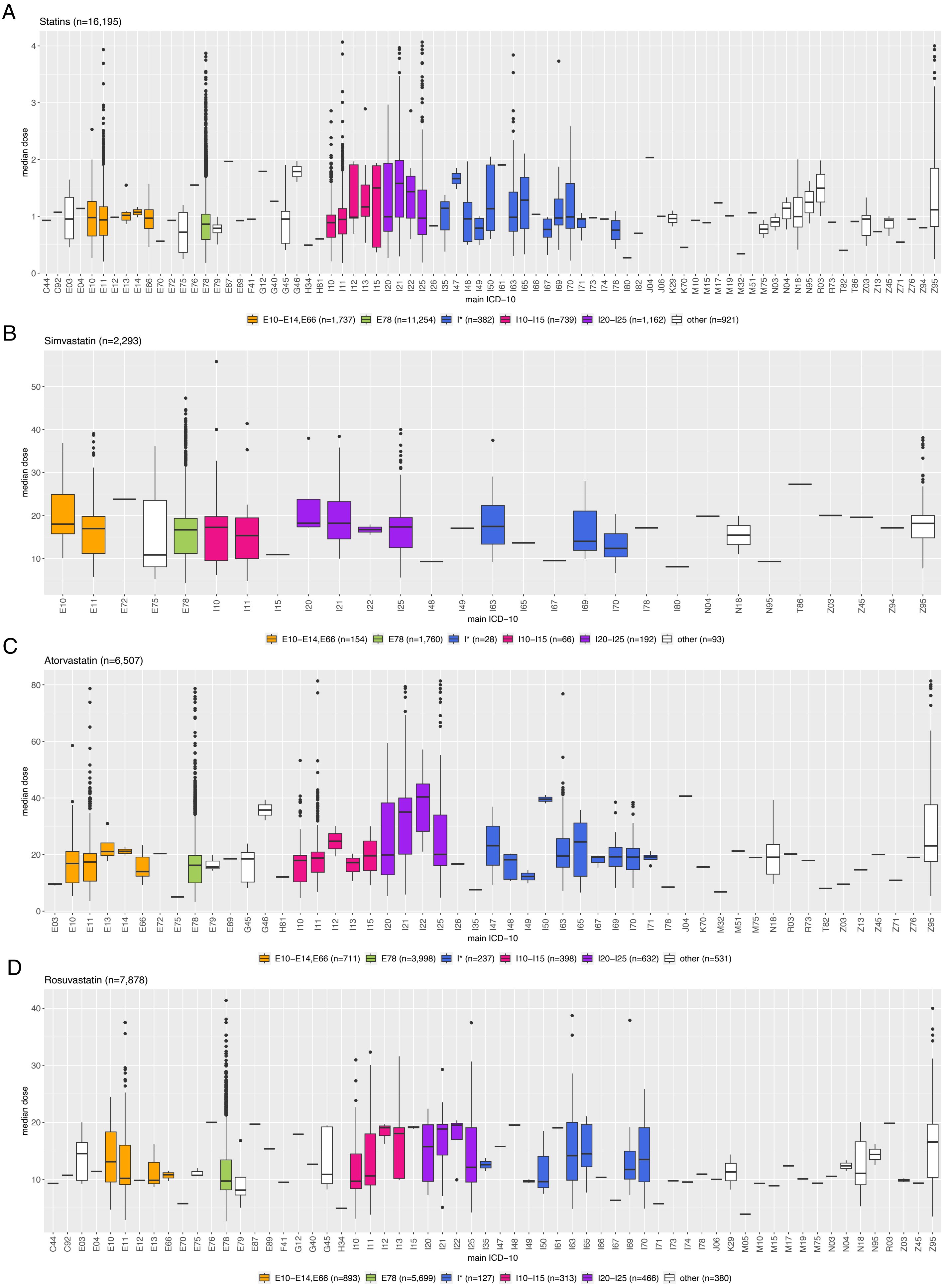
A
Statins (n=16,195)
median dose
main ICD−10
E10−E14,E66 (n=1,737) E78 (n=11,254) I* (n=382) I10−I15 (n=739) I20−I25 (n=1,162) other (n=921)
B
Simvastatin (n=2,293)
median dose
main ICD−10
E10−E14,E66 (n=154) E78 (n=1,760) I* (n=28) I10−I15 (n=66) I20−I25 (n=192) other (n=93)
C
Atorvastatin (n=6,507)
median dose
main ICD−10
E10−E14,E66 (n=711) E78 (n=3,998) I* (n=237) I10−I15 (n=398) I20−I25 (n=632) other (n=531)
D
Rosuvastatin (n=7,878)
median dose
main ICD−10
E10−E14,E66 (n=893) E78 (n=5,699) I* (n=127) I10−I15 (n=313) I20−I25 (n=466) other (n=380)

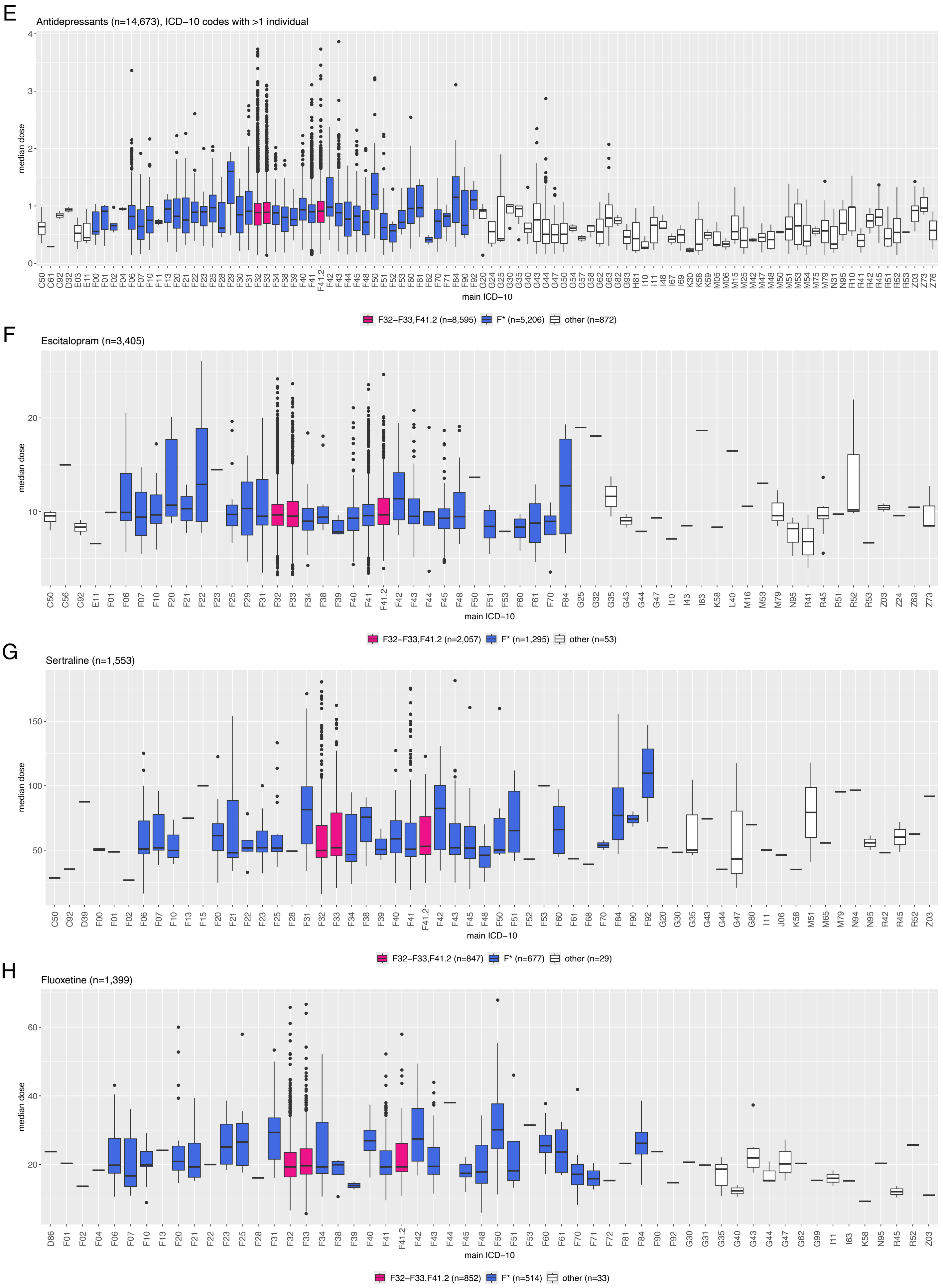
E
Antidepressants (n=14,673), ICD−10 codes with >1 individual
median dose
main ICD−10
F32−F33,F41.2 (n=8,595)
F* (n=5,206)
other (n=872)
F
Escitalopram (n=3,405)
median dose
main ICD−10
F32−F33,F41.2 (n=2,057)
F* (n=1,295)
other (n=53)
G
Sertraline (n=1,553)
median dose
main ICD−10
F32−F33,F41.2 (n=847)
F* (n=677)
other (n=29)
H
Fluoxetine (n=1,399)
median dose
main ICD−10
F32−F33,F41.2 (n=852)
F* (n=514)
other (n=33)

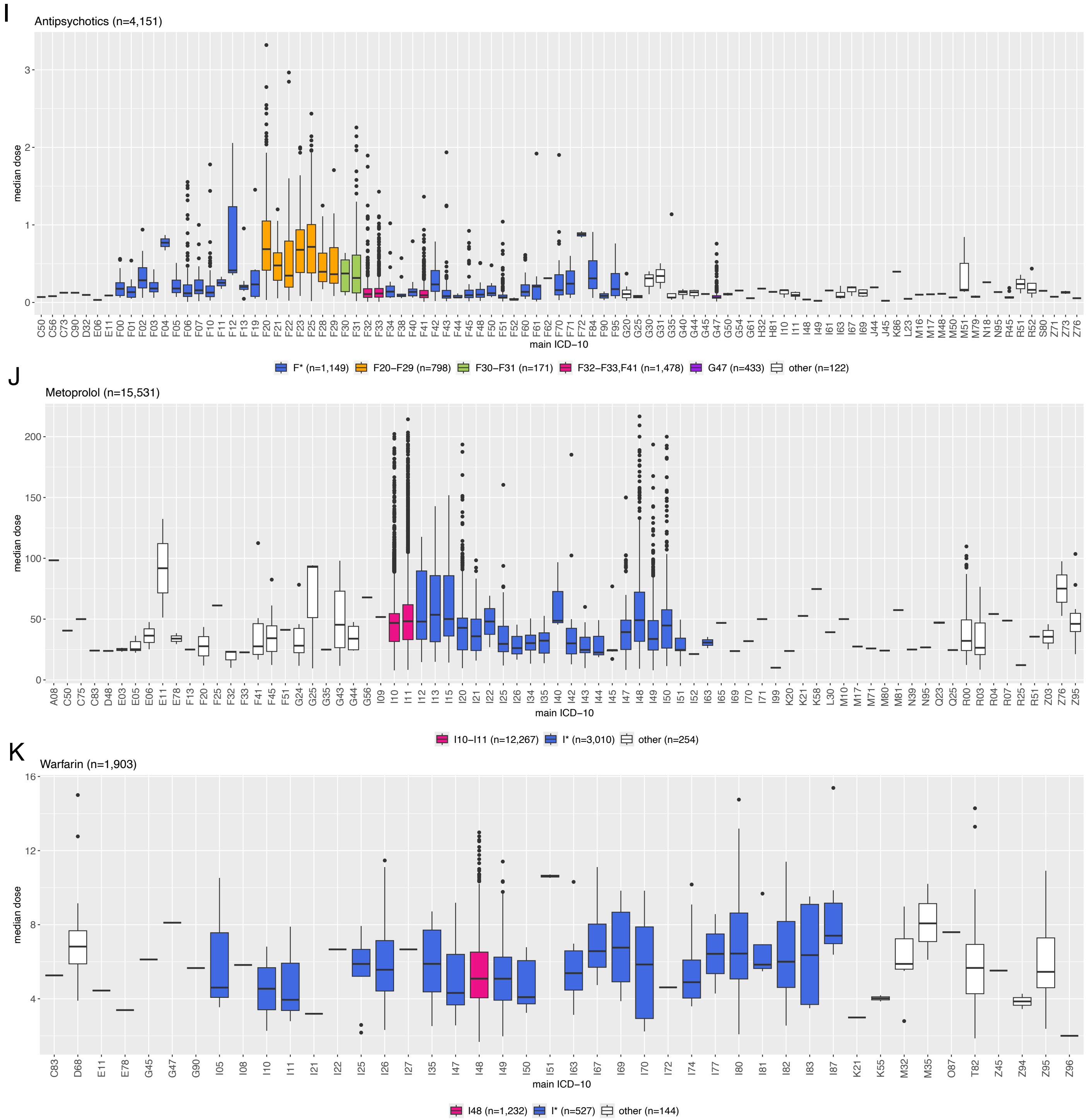

**Supplementary Figure 7. Overview of the distribution of derived median doses across purchases by the primary ICD-10 on prescription.** Median dose is reported in milligrams for individual drugs and standardized to DDD for class-level medications. The distributions are outlined for (A) statins, (B) simvastatin, (C) atorvastatin, (D) rosuvastatin, (E) antidepressants, (F) escitalopram, (G) sertraline, (H) fluoxetine, (I) antipsychotics, (J) metoprolol, and (K) warfarin. ICD-10 codes are colour-coded by diagnostic groups to highlight the most prevalent ICD-10 codes and distinct endpoints by drug. Specifically, for statins, simvastatin, atorvastatin, and rosuvastatin: E10-E14, E66 – diabetes, obesity; E78 – hypercholesterolemia; I10-I15 – hypertension; I20-I25 – coronary heart disease; I* – diseases of the circulatory system. For antidepressants, escitalopram, sertraline, and fluoxetine: F32-F33, F41.2 – depression; F* – mental and behavioural disorders. For antipsychotics: F20-F29 – schizophrenia spectrum disorder; F30-F31 – mania, bipolar disorder; F32-F33, F41 – depression, anxiety; G47 – sleep disorders; F* – mental and behavioural disorders. For metoprolol: I10-I11 – essential hypertension and hypertensive heart disease; I* – diseases of the circulatory system. For warfarin: I48 – atrial fibrillation and flutter; I* – diseases of the circulatory system. Of note, the sample sizes for individuals taking antidepressants, escitalopram, sertraline, and fluoxetine with F32, F33, F41.2 on prescription (E-H) and individuals taking antipsychotics with F20-F29 on prescriptions (I) in the figure differ from the sample set used in association testing (Table 1). This discrepancy stems from the 3SD filter (exclusion of doses deviating >3SDs from the log-scale mean), and the related sample exclusion (exclusion of one individual per related pair). These filters were applied across all drug users (in figure) or across subgroup (for association testing to maximize the number of cases).

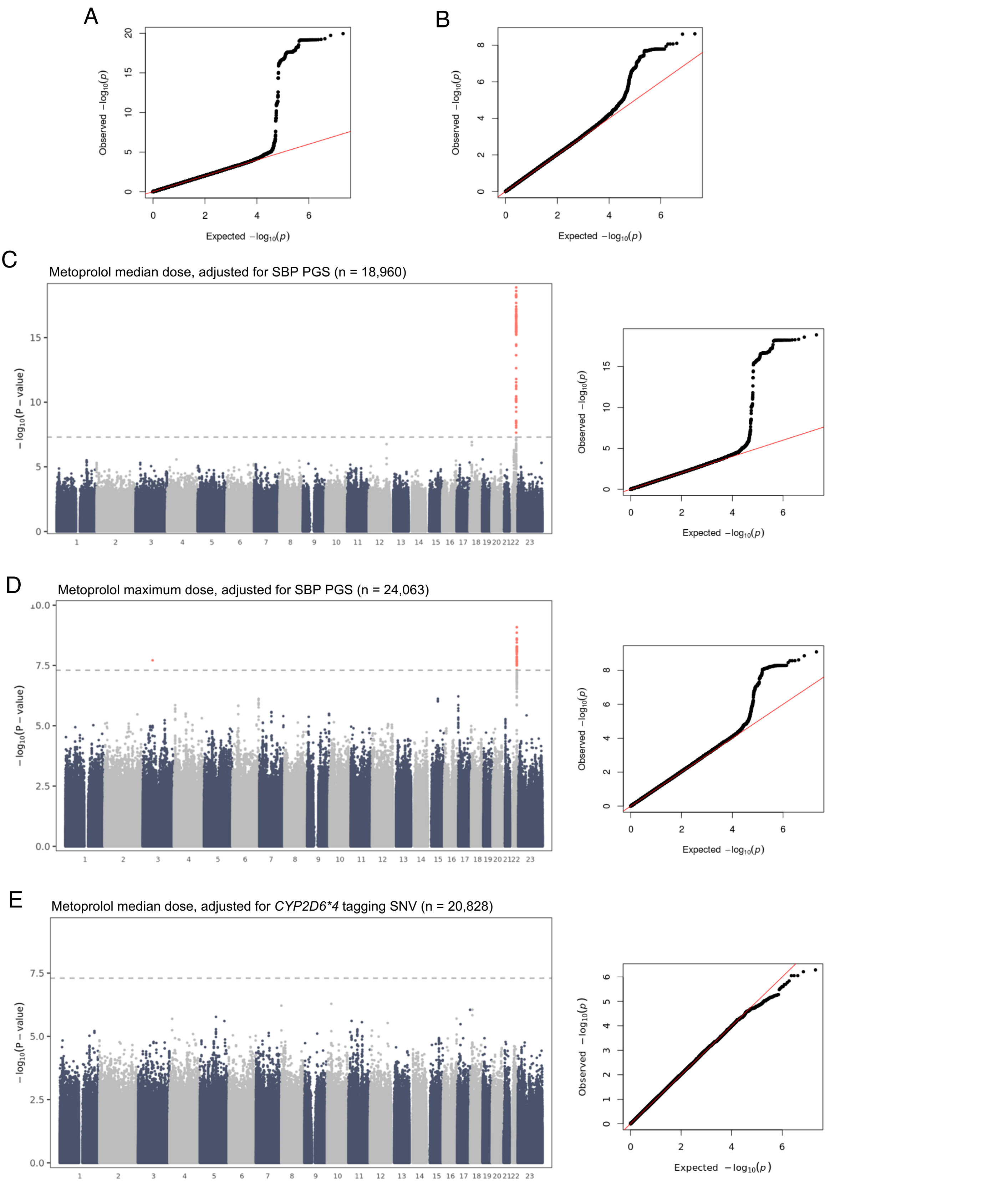

**Supplementary Figure 8. GWAS results for metoprolol median dose and maximum dose.** (A-B) QQ plots for (A) median and (B) maximum dose results. (C-D) Manhattan and QQ plots for (C) median and (D) maximum dose results, adjusted for SBP PGS. (E) Manhattan and QQ plot for median dose results, adjusted for *CYP2D6*4* tag-SNV. Genome-wide significance ($P<5\times10^{-8}$) is shown as a dashed line, genome-wide significant variants are highlighted in red, and number 23 on x-axis denotes chromosome X. Lambda values: (A) 1.02, (B) 1.03, (C) 1.02, (D) 1.03, (E) 1.02.

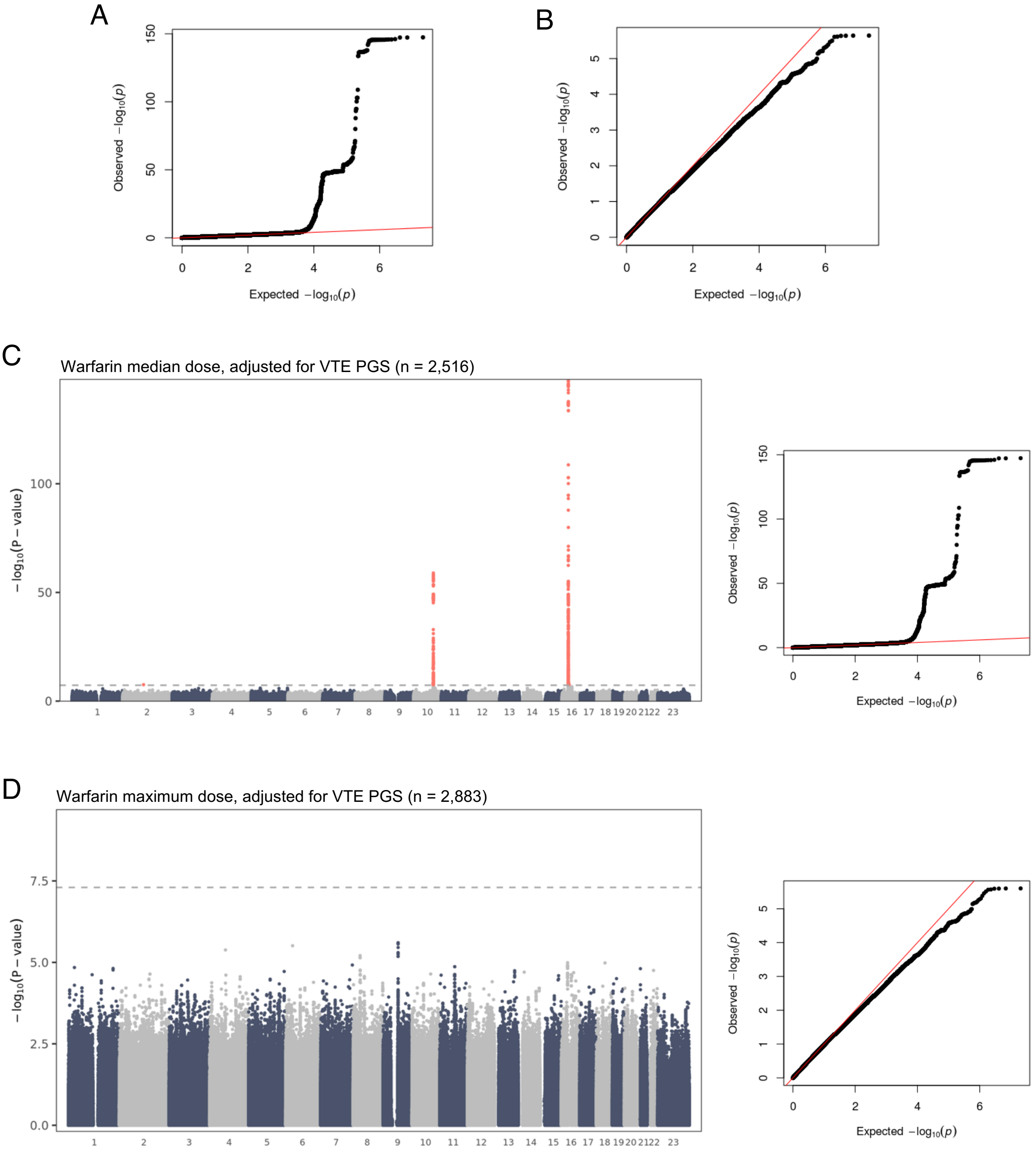

**Supplementary Figure 9. GWAS results for warfarin median dose and maximum dose.** (A-B) QQ plots for (A) median and (B) maximum dose results. (C-D) Manhattan and QQ plots for (C) median and (D) maximum dose results, adjusted for VTE PGS. Genome-wide significance ($P<5\times10^{-8}$) is shown as a dashed line, genome-wide significant variants are highlighted in red, and number 23 on x-axis denotes chromosome X. Lambda values: (A) 1.01, (B) 0.99, (C) 1.01, (D) 0.99.

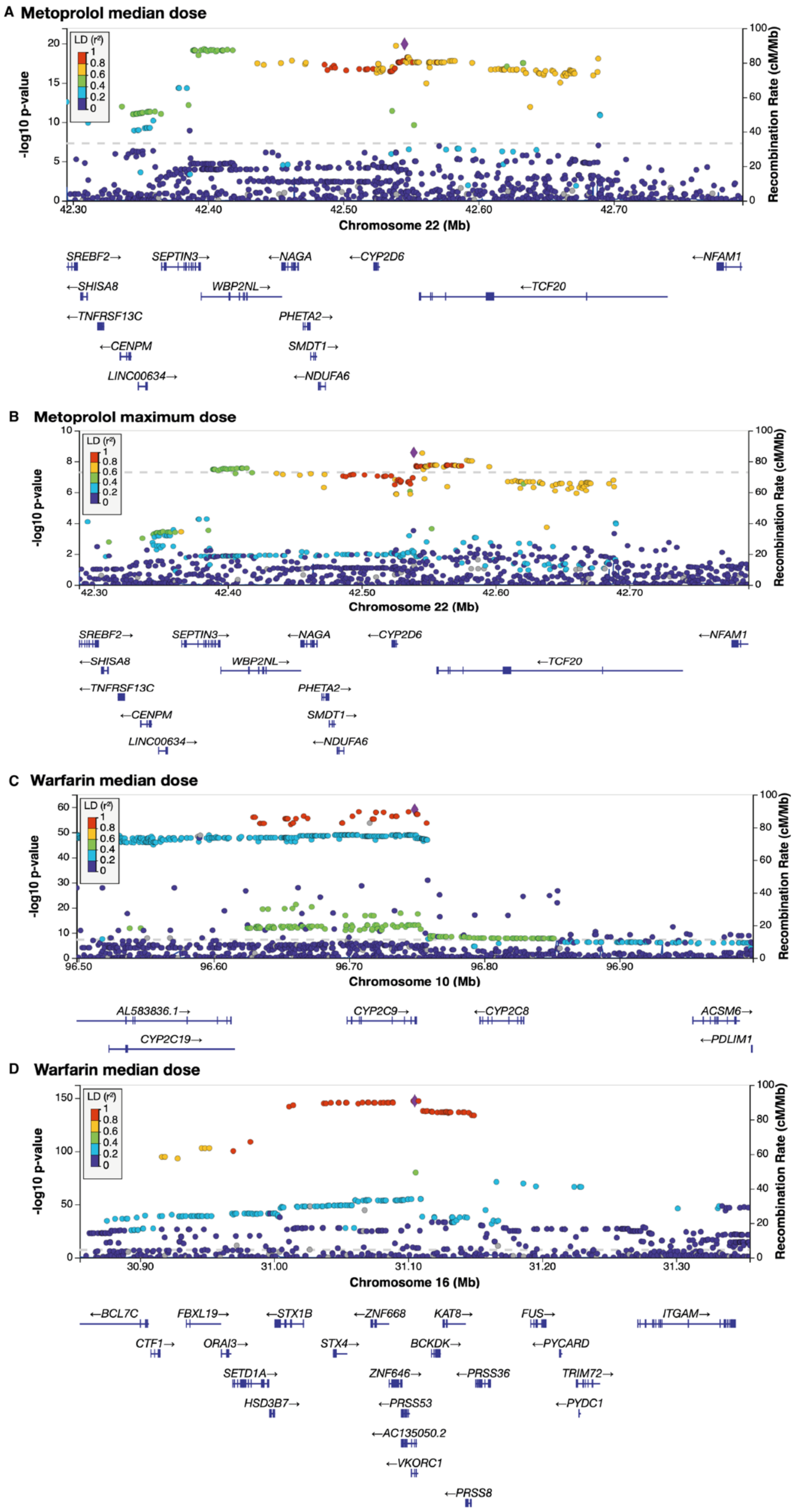

**Supplementary Figure 10. The regional association plots for metoprolol median and maximum dose (A,B) and warfarin median dose (C,D).** The y-axis represents the statistical significance (–$\log_{10}$(P-value)), the x-axis genomic position (Mb) with gene coordinates according to GENCODE GRCh37 in the UCSC Genome Browser, and z-axis the recombination rate (cM/Mb). A blue line denotes the recombination rate. The purple diamond marks the most significant SNV within each locus. SNVs are colored-coded based on LD ($r^2$) with the lead SNV, calculated using the European (EUR) population reference from the 1000 Genomes Project. The strongest signal (rs5751229) for metoprolol median dose is located 18 kb from the transcription start site of *CYP2D6* (A). Because this exceeds the 5 kb threshold applied for listing the top five SNVs per PGx gene in Supplementary Table 9, it is not included in that table.

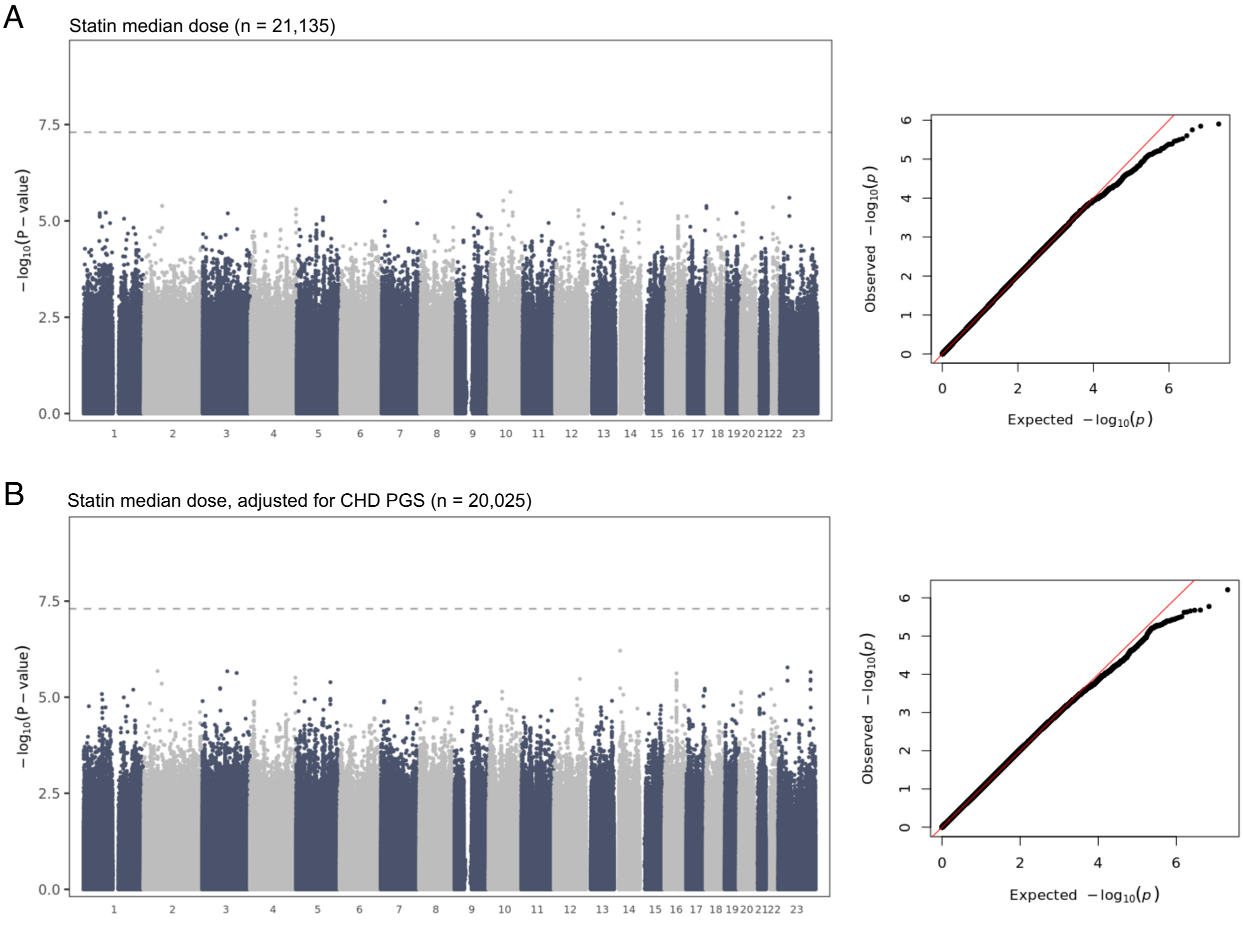

**Supplementary Figure 11. GWAS results for statin median dose.** (A-B) Manhattan and QQ plots for median dose (A) without and (B) with CHD PGS in the model. Genome-wide significance ($P<5\times10^{-8}$) is shown as a dashed line. Genome-wide significance ($P<5\times10^{-8}$) is shown as a dashed line, and number 23 on x-axis denotes chromosome X. Lambda values: (A) 1.02, (B) 1.03.

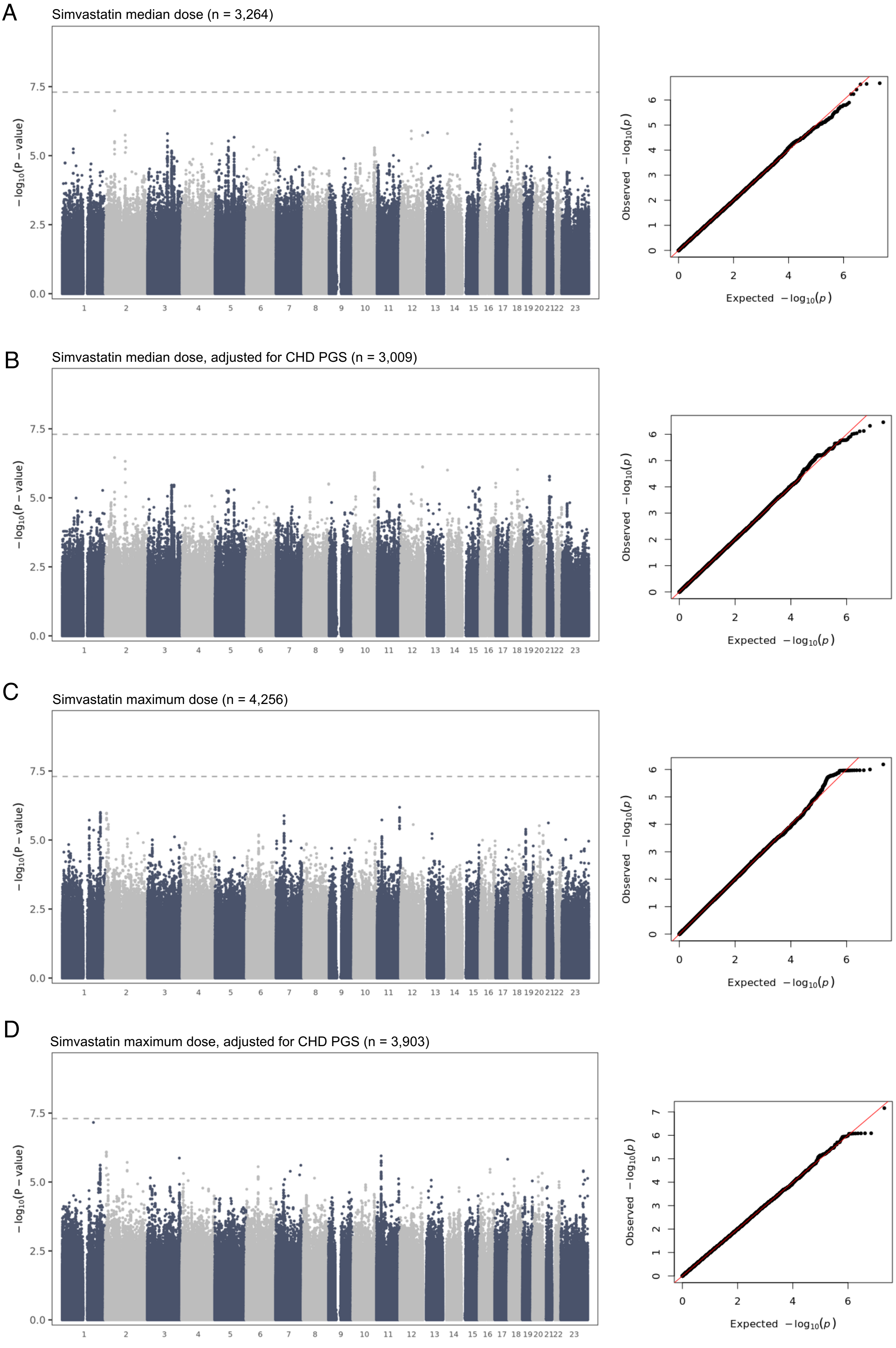

**Supplementary Figure 12. GWAS results for simvastatin median dose and maximum dose.** (A-B) Manhattan and QQ plots for median dose (A) without and (B) with CHD PGS in the model. (C-D) Manhattan and QQ plots for maximum dose (C) without and (D) with CHD PGS in the model. Genome-wide significance ($P<5\times10^{-8}$) is shown as a dashed line, and number 23 on x-axis denotes chromosome X. Lambda values: (A) 0.99, (B) 0.99, (C) 1.02, (D) 1.01.

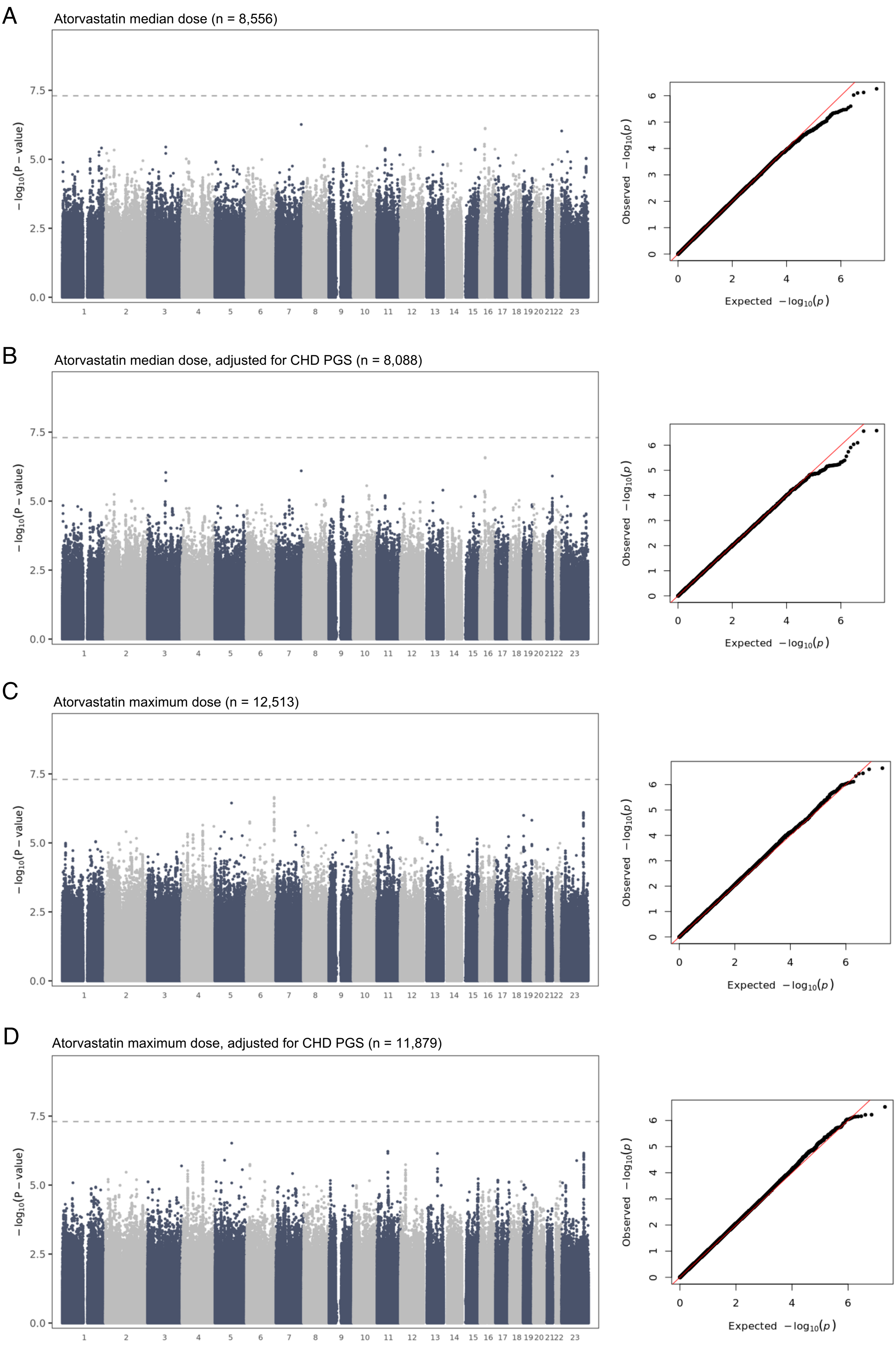

**Supplementary Figure 13. GWAS results for atorvastatin median dose and maximum dose.** (A-B) Manhattan and QQ plots for median dose (A) without and (B) with CHD PGS in the model. (C-D) Manhattan and QQ plots for maximum dose (C) without and (D) with CHD PGS in the model. Genome-wide significance ($P<5\times10^{-8}$) is shown as a dashed line, and number 23 on x-axis denotes chromosome X. Lambda values: (A) 1.01, (B) 1.01, (C) 1.05, (D) 1.04.

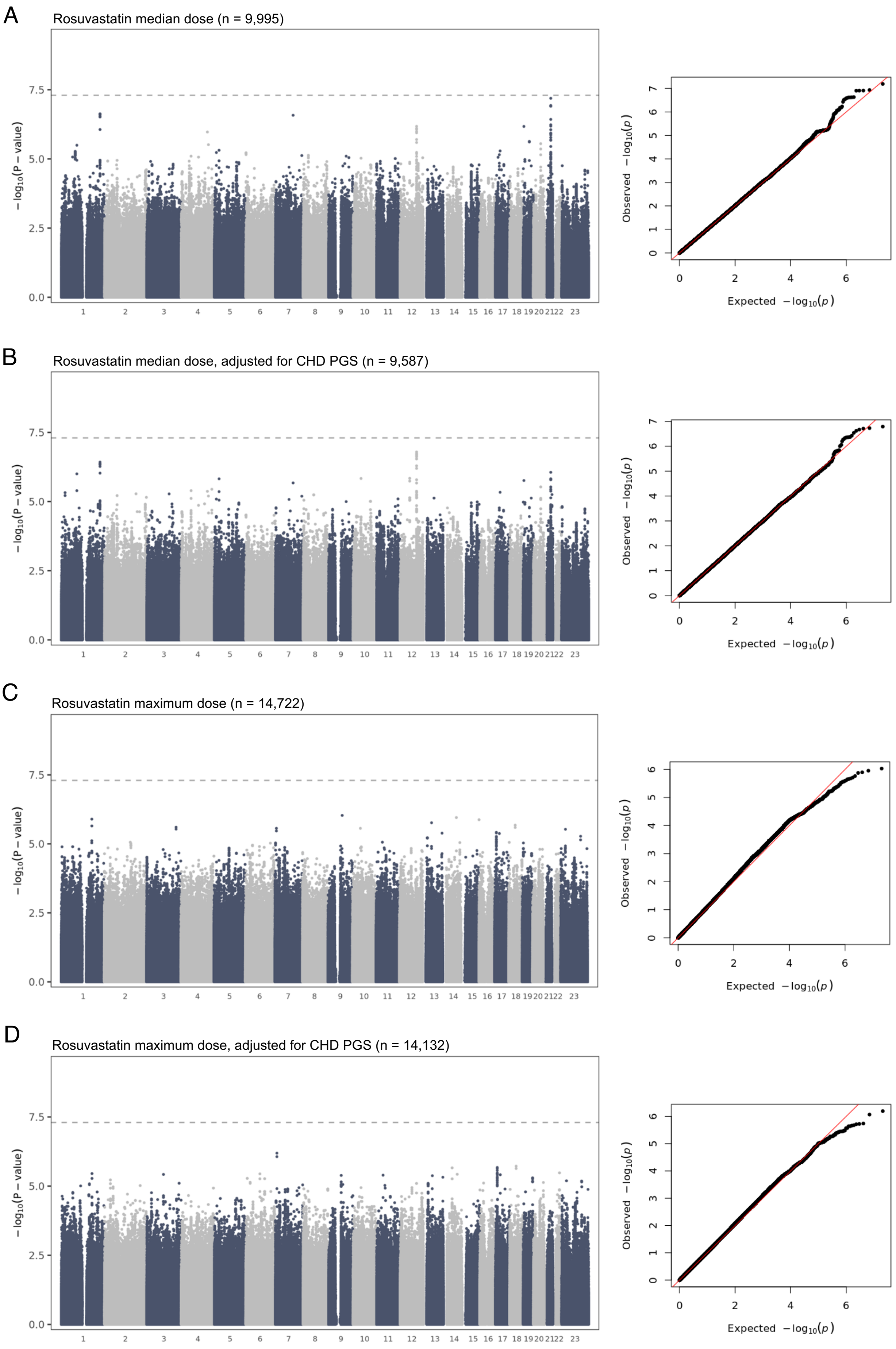

**Supplementary Figure 14. GWAS results for rosuvastatin median dose and maximum dose.** (A-B) Manhattan and QQ plots for median dose (A) without and (B) with CHD PGS in the model. (C-D) Manhattan and QQ plots for maximum dose (C) without and (D) with CHD PGS in the model. Genome-wide significance ($P<5\times10^{-8}$) is shown as a dashed line, and number 23 on x-axis denotes chromosome X. Lambda values: (A) 1.00, (B) 0.99, (C) 1.06, (D) 1.03.

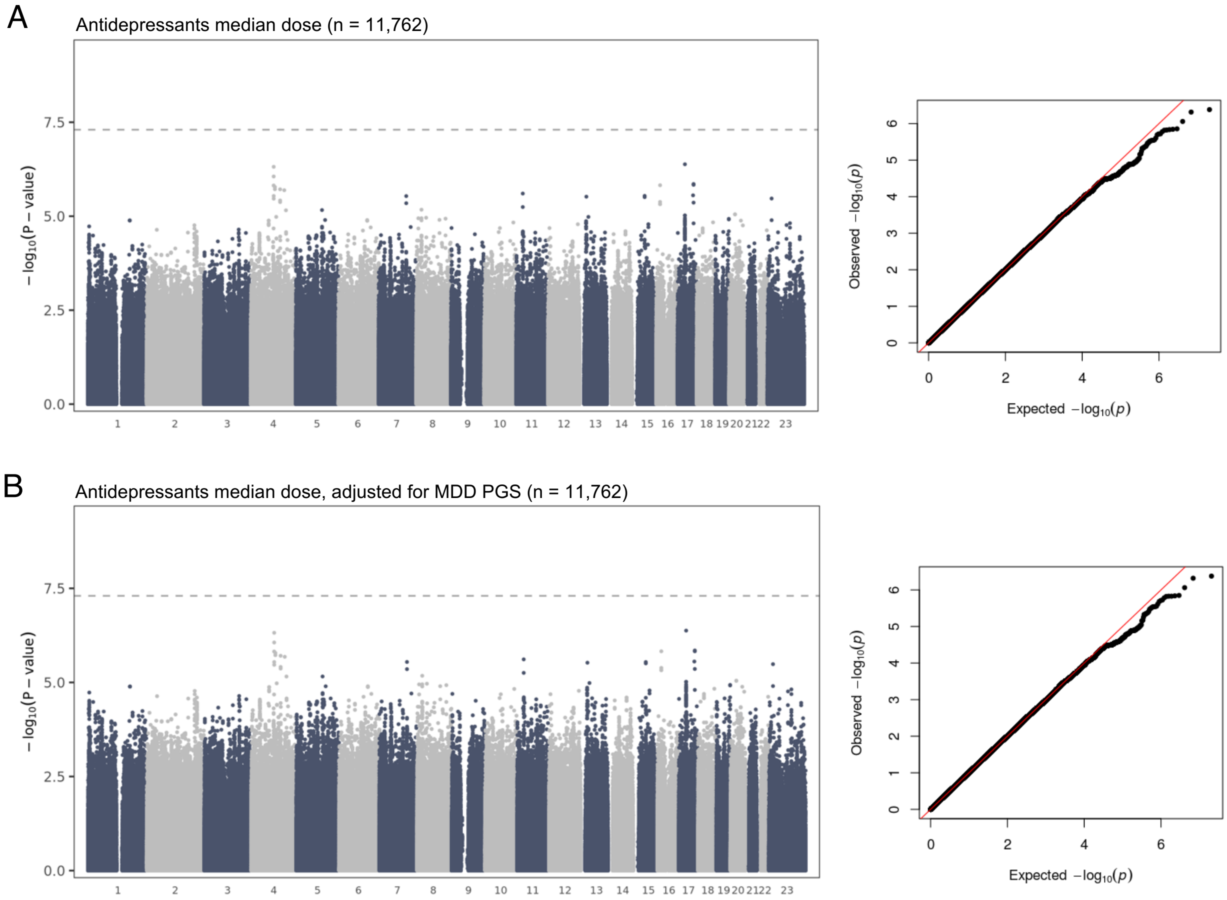

**Supplementary Figure 15. GWAS results for antidepressant median dose.** (A-B) Manhattan and QQ plots for median dose (A) without and (B) with MDD PGS in the model. Genome-wide significance ($P<5\times10^{-8}$) is shown as a dashed line, and number 23 on x-axis denotes chromosome X. Lambda values: (A) 1.01, (B) 1.01.

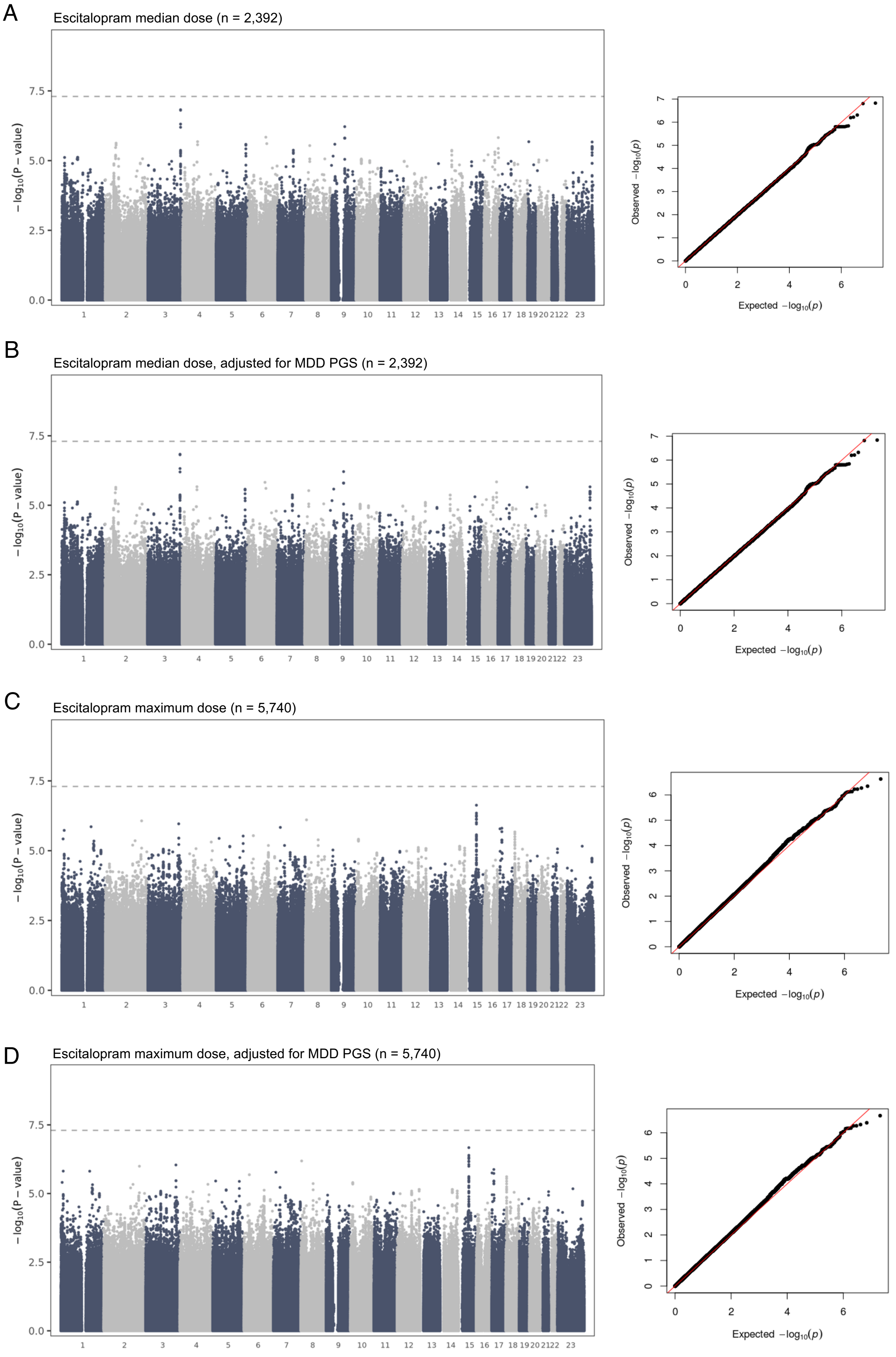

**Supplementary Figure 16. GWAS results for escitalopram median dose and maximum dose.** (A-B) Manhattan and QQ plots for median dose (A) without and (B) with MDD PGS in the model. (C-D) Manhattan and QQ plots for maximum dose (C) without and (D) with MDD PGS in the model. Genome-wide significance ($P<5\times10^{-8}$) is shown as a dashed line, and number 23 on x-axis denotes chromosome X. Lambda values: (A) 0.99, (B) 0.99, (C) 1.05, (D) 1.05.

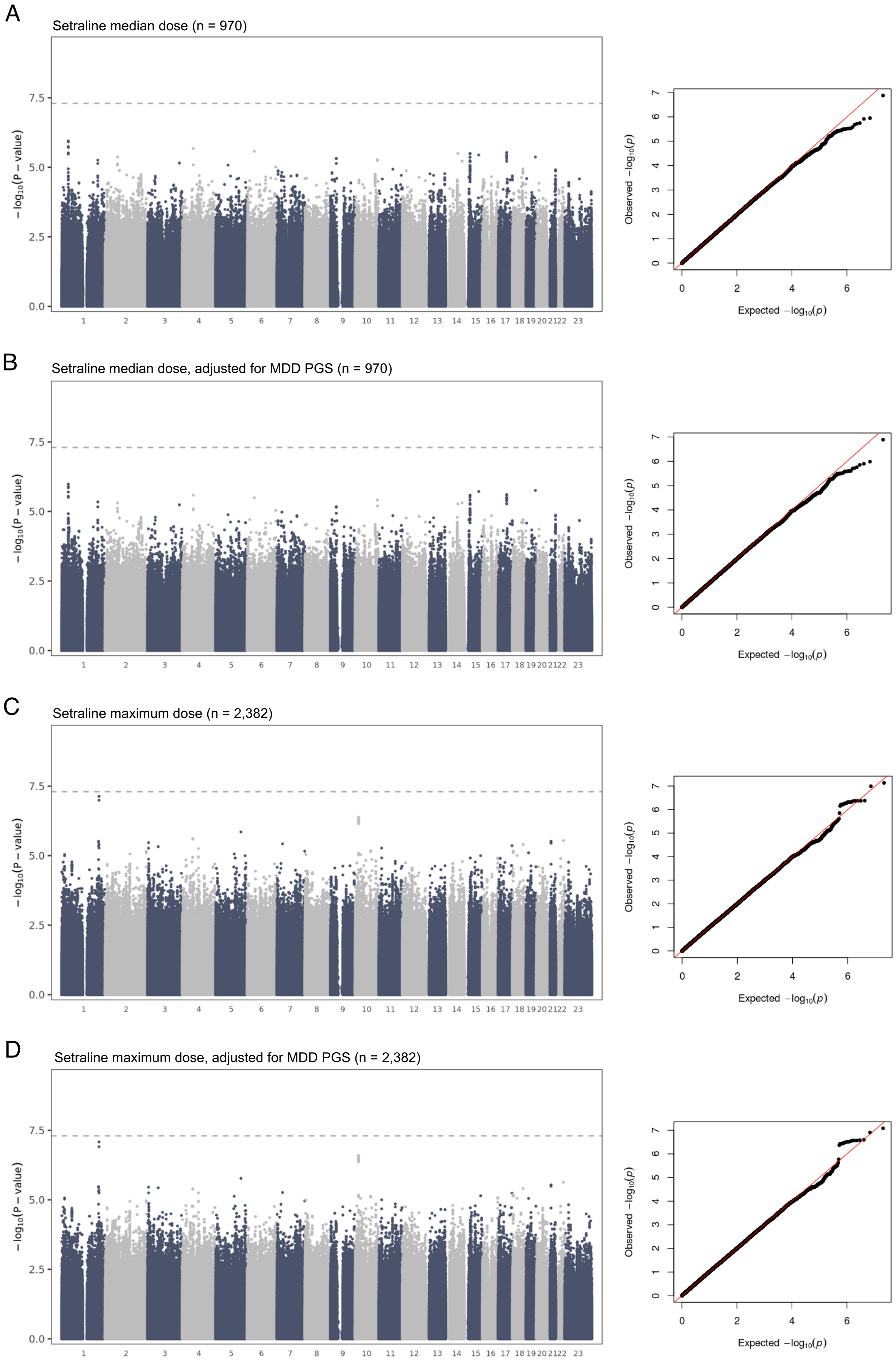

**Supplementary Figure 17. GWAS results for sertraline median dose and maximum dose.** (A-B) Manhattan and QQ plots for median dose (A) without and (B) with MDD PGS in the. model. (C-D) Manhattan and QQ plots for maximum dose (C) without and (D) with MDD PGS in the model. Genome-wide significance ($P<5\times10^{-8}$) is shown as a dashed line, and number 23 on x-axis denotes chromosome X. Lambda values: (A) 0.99, (B) 0.99, (C) 1.03, (D) 1.03.

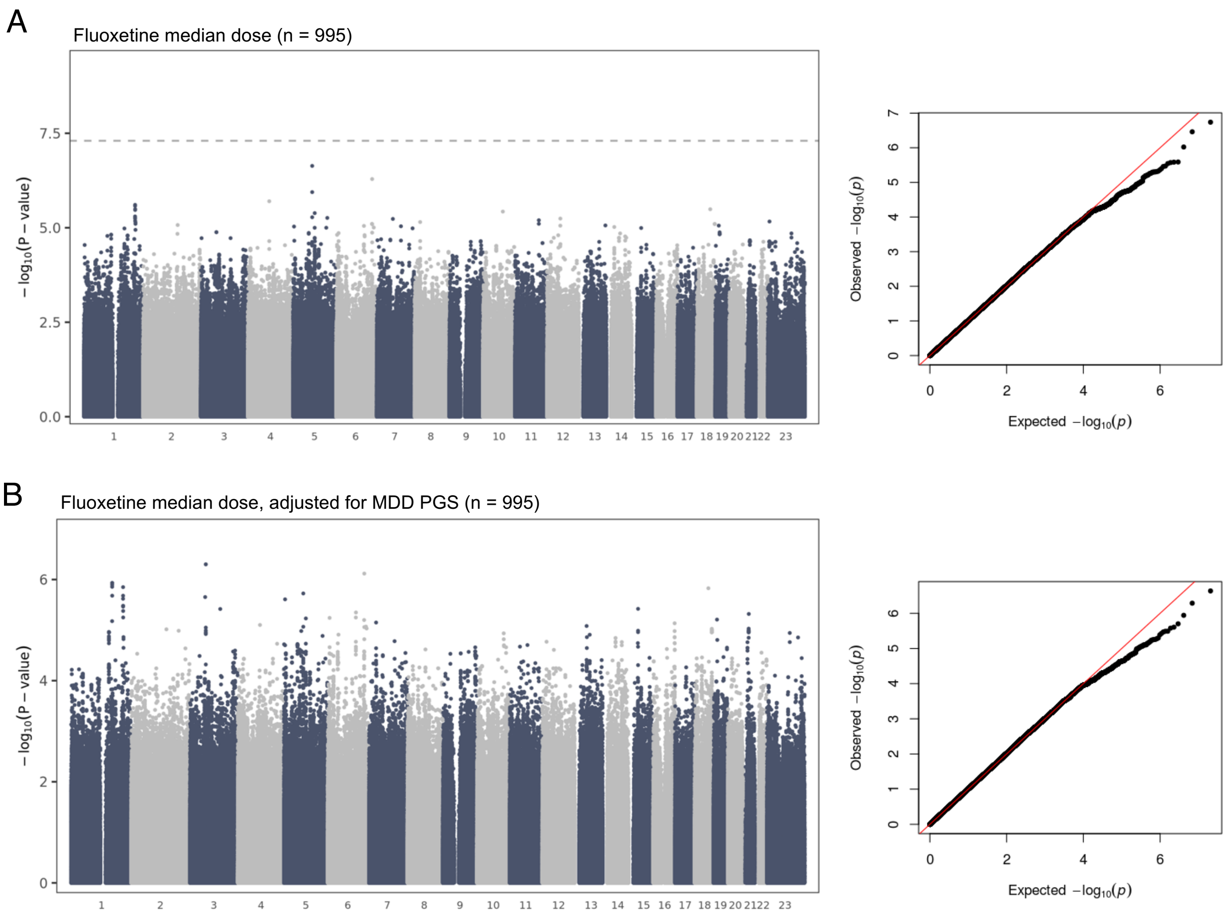

**Supplementary Figure 18. GWAS results for fluoxetine median dose.** (A-B) Manhattan and QQ plots for median dose (A) without and (B) with MDD PGS in the model. Genome-wide significance ($P<5\times10^{-8}$) is shown as a dashed line, and number 23 on x-axis denotes chromosome X. Lambda values: (A) 1.01, (B) 1.01, (C) 1.06, (D) 1.03.

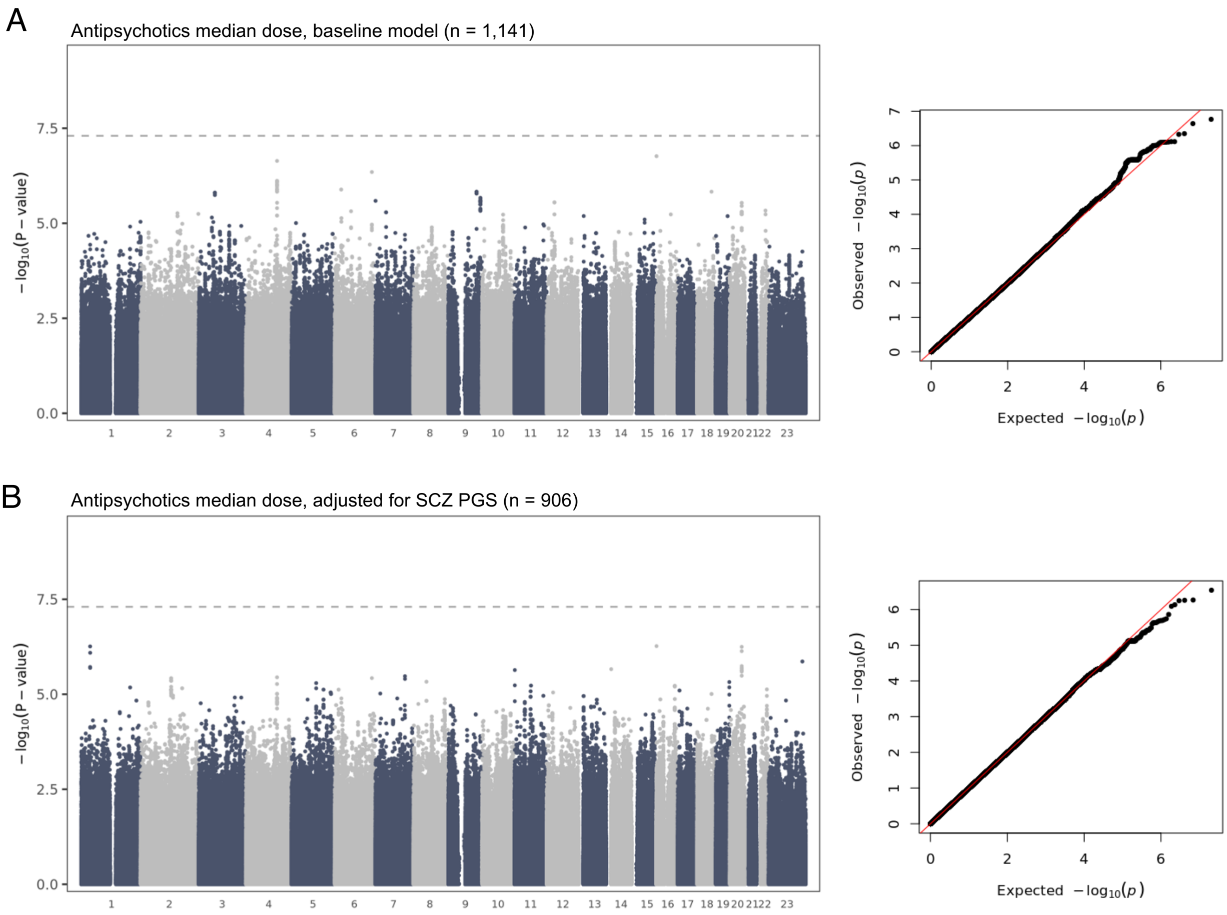

**Supplementary Figure 19. GWAS results for antipsychotic median dose.** (A-B) Manhattan and QQ plots for median dose (A) without and (B) with SCZ PGS in the model. Genome-wide significance ($P<5\times10^{-8}$) is shown as a dashed line, and number 23 on x-axis denotes chromosome X. Lambda values: (A) 0.99, (B) 0.99.

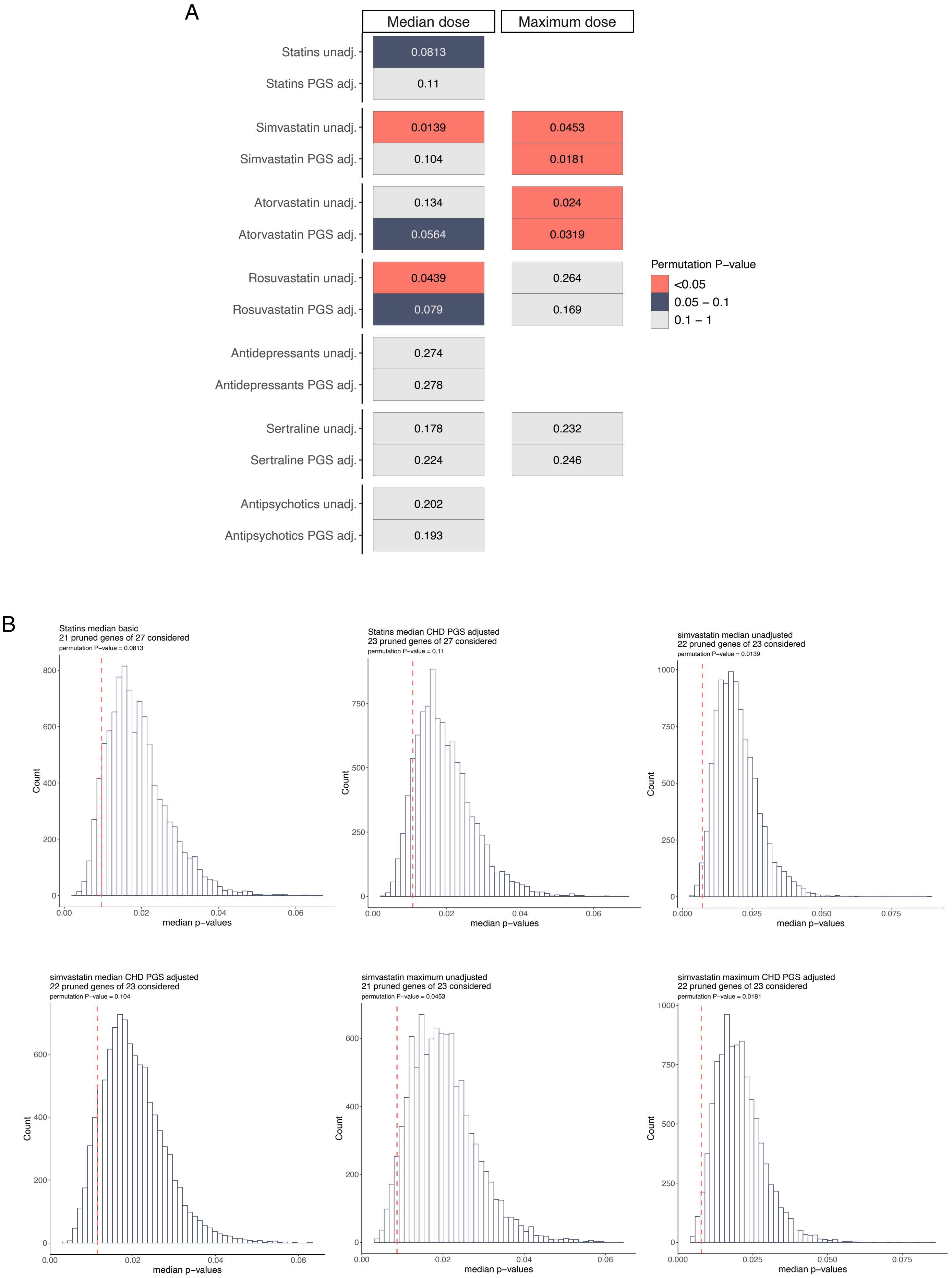
A
Median dose
Maximum dose
Statins unadj.
Statins PGS adj.
Simvastatin unadj.
Simvastatin PGS adj.
Atorvastatin unadj.
Atorvastatin PGS adj.
Rosuvastatin unadj.
Rosuvastatin PGS adj.
Antidepressants unadj.
Antidepressants PGS adj.
Sertraline unadj.
Sertraline PGS adj.
Antipsychotics unadj.
Antipsychotics PGS adj.
0.0813
0.11
0.0139
0.0453
0.104
0.0181
0.134
0.024
0.0564
0.0319
0.0439
0.264
0.079
0.169
0.274
0.278
0.178
0.232
0.224
0.246
0.202
0.193
Permutation P−value
<0.05
0.05 – 0.1
0.1 – 1
B
Statins median basic
21 pruned genes of 27 considered
Statins median CHD PGS adjusted
23 pruned genes of 27 considered
simvastatin median unadjusted
22 pruned genes of 23 considered
simvastatin median CHD PGS adjusted
22 pruned genes of 23 considered
simvastatin maximum unadjusted
21 pruned genes of 23 considered
simvastatin maximum CHD PGS adjusted
22 pruned genes of 23 considered
Count
median p−values

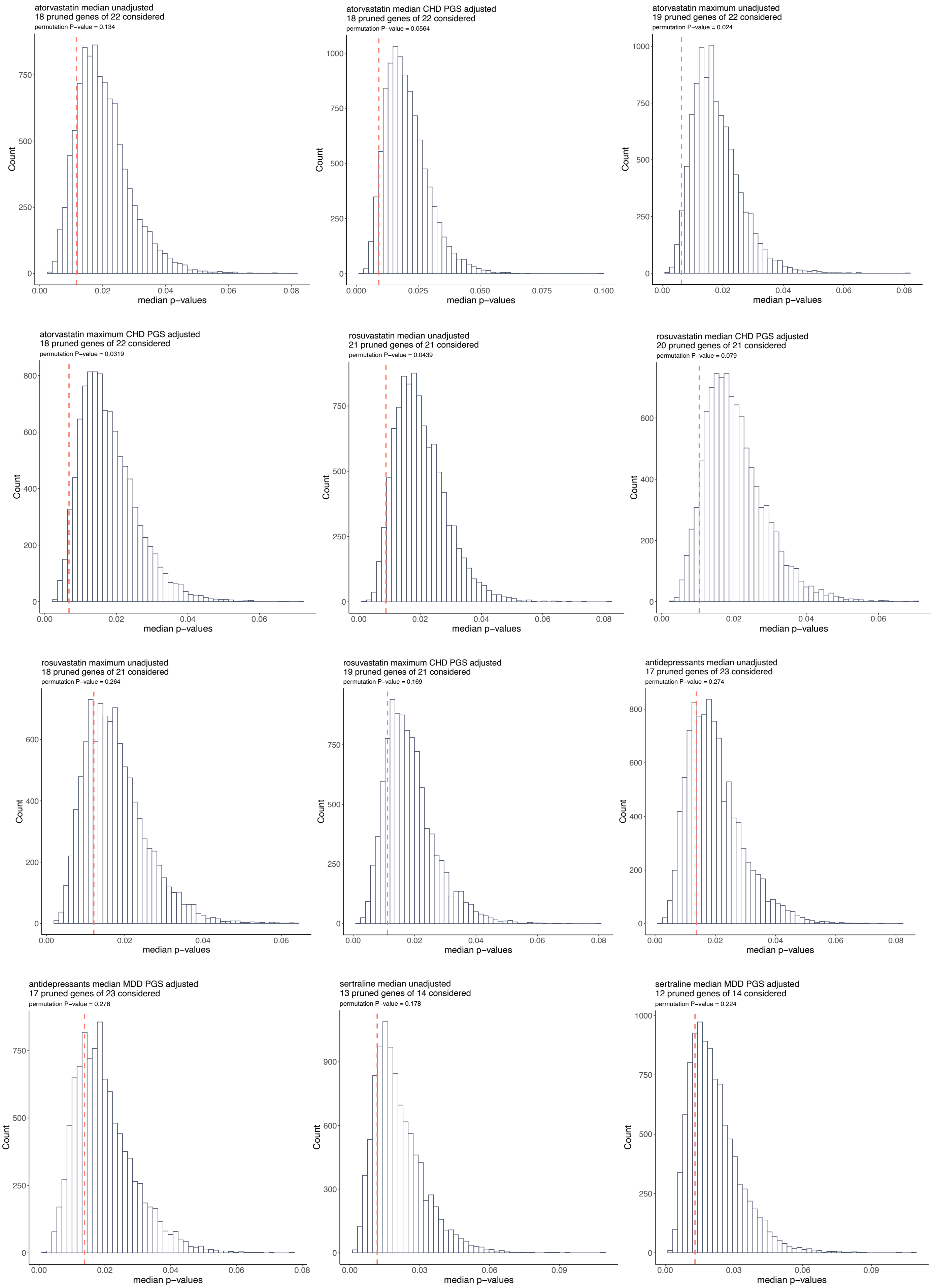
atorvastatin median unadjusted
18 pruned genes of 22 considered
permutation P−value = 0.134
atorvastatin median CHD PGS adjusted
18 pruned genes of 22 considered
permutation P−value = 0.0564
atorvastatin maximum unadjusted
19 pruned genes of 22 considered
permutation P−value = 0.024
atorvastatin maximum CHD PGS adjusted
18 pruned genes of 22 considered
permutation P−value = 0.0319
rosuvastatin median unadjusted
21 pruned genes of 21 considered
permutation P−value = 0.0439
rosuvastatin median CHD PGS adjusted
20 pruned genes of 21 considered
permutation P−value = 0.079
rosuvastatin maximum unadjusted
18 pruned genes of 21 considered
permutation P−value = 0.264
rosuvastatin maximum CHD PGS adjusted
19 pruned genes of 21 considered
permutation P−value = 0.169
antidepressants median unadjusted
17 pruned genes of 23 considered
permutation P−value = 0.274
antidepressants median MDD PGS adjusted
17 pruned genes of 23 considered
permutation P−value = 0.278
sertraline median unadjusted
13 pruned genes of 14 considered
permutation P−value = 0.178
sertraline median MDD PGS adjusted
12 pruned genes of 14 considered
permutation P−value = 0.224
Count
median p−values

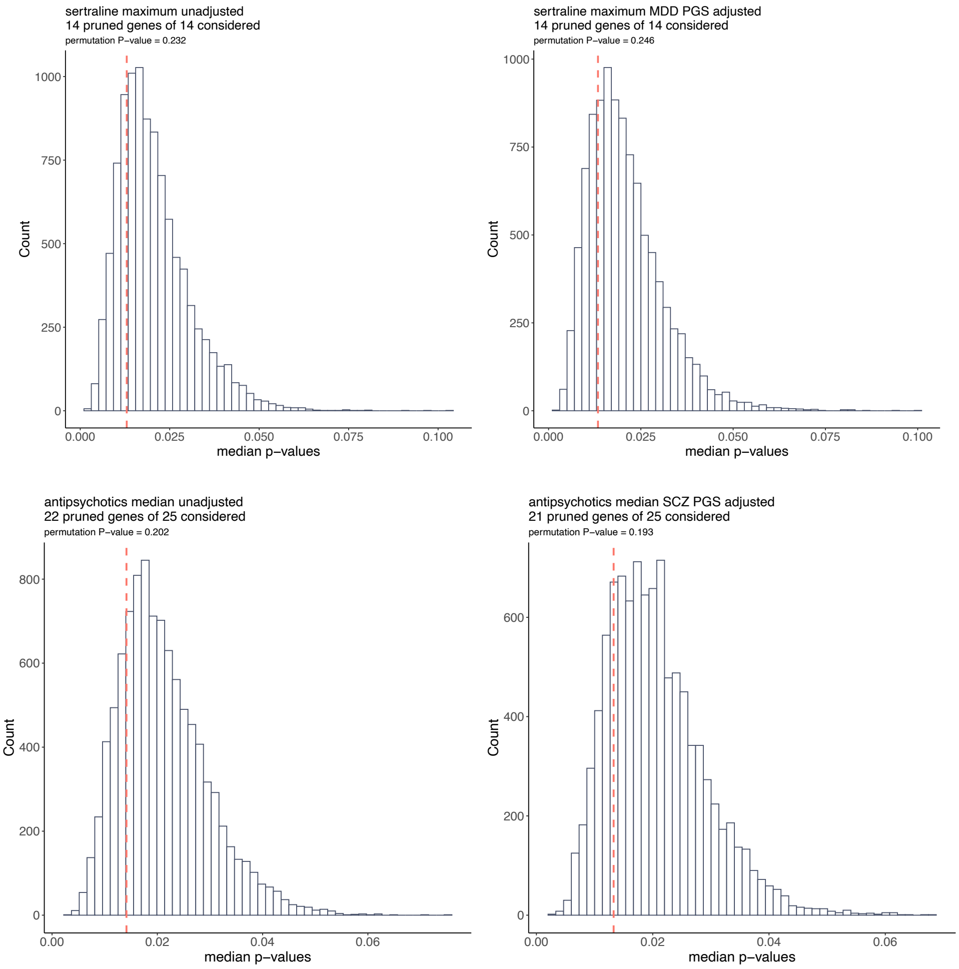

**Supplementary Figure 20. Permutation-based evaluation of PGx gene associations in GWAS results.** (A) Permutation p-values testing whether drug-specific PGx genes show stronger associations than expected by chance in GWAS of median and maximum dose for statins, antidepressants, and antipsychotics, with and without CHD PGS adjustment. The target trait and test are outlined on y-axis with the dose metric separated into two panels (median dose or maximum dose). P-values are colour-coded according to significance thresholds. (B) Permutation-based null distribution of median p-values for drug-specific PGx gene sets in GWAS results. For each permutation, a median p-value was calculated from an equal number of randomly sampled genes from a background set, generating the null distribution. The observed median p-value (red dashed line) was then compared to this distribution to determine whether PGx gene associations deviated from expectation. The permutation p-value represents the proportion of permuted median p-values that were as extreme or lower than the observed value.

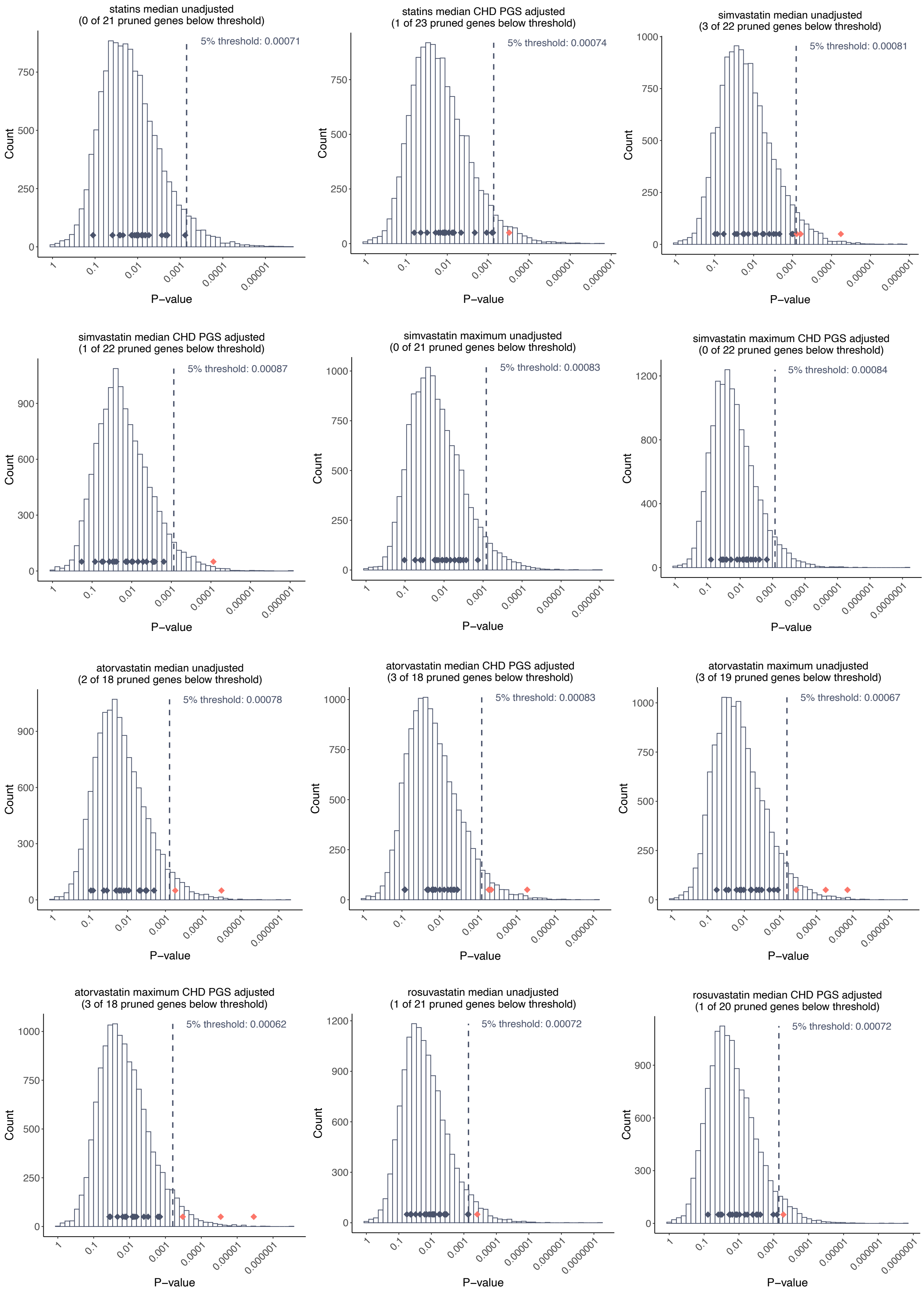
statins median unadjusted
(0 of 21 pruned genes below threshold)
5% threshold: 0.00071
statins median CHD PGS adjusted
(1 of 23 pruned genes below threshold)
5% threshold: 0.00074
simvastatin median unadjusted
(3 of 22 pruned genes below threshold)
5% threshold: 0.00081
simvastatin median CHD PGS adjusted
(1 of 22 pruned genes below threshold)
5% threshold: 0.00087
simvastatin maximum unadjusted
(0 of 21 pruned genes below threshold)
5% threshold: 0.00083
simvastatin maximum CHD PGS adjusted
(0 of 22 pruned genes below threshold)
5% threshold: 0.00084
atorvastatin median unadjusted
(2 of 18 pruned genes below threshold)
5% threshold: 0.00078
atorvastatin median CHD PGS adjusted
(3 of 18 pruned genes below threshold)
5% threshold: 0.00083
atorvastatin maximum unadjusted
(3 of 19 pruned genes below threshold)
5% threshold: 0.00067
atorvastatin maximum CHD PGS adjusted
(3 of 18 pruned genes below threshold)
5% threshold: 0.00062
rosuvastatin median unadjusted
(1 of 21 pruned genes below threshold)
5% threshold: 0.00072
rosuvastatin median CHD PGS adjusted
(1 of 20 pruned genes below threshold)
5% threshold: 0.00072
Count
P−value

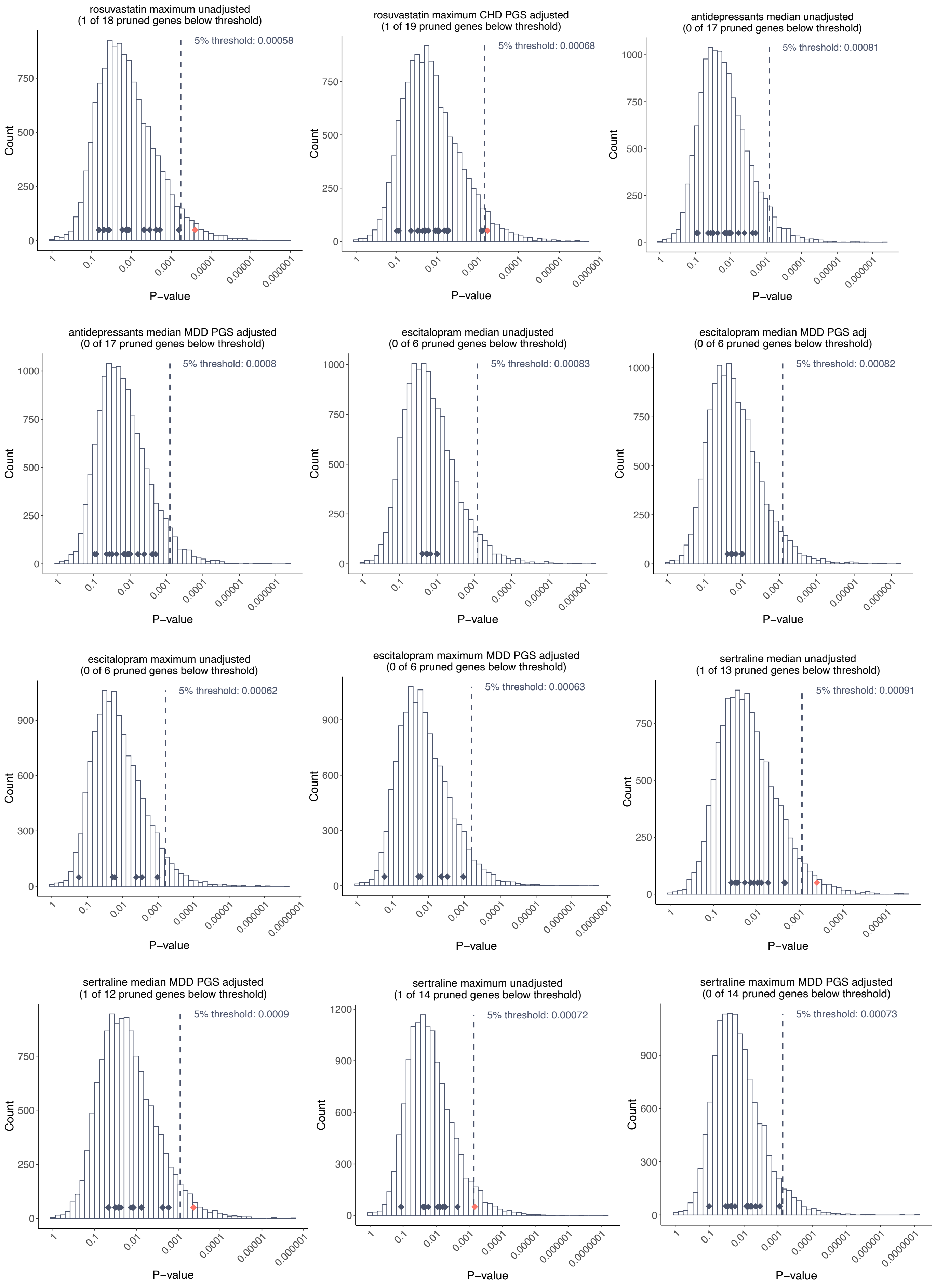
rosuvastatin maximum unadjusted
(1 of 18 pruned genes below threshold)
5% threshold: 0.00058
rosuvastatin maximum CHD PGS adjusted
(1 of 19 pruned genes below threshold)
5% threshold: 0.00068
antidepressants median unadjusted
(0 of 17 pruned genes below threshold)
5% threshold: 0.00081
antidepressants median MDD PGS adjusted
(0 of 17 pruned genes below threshold)
5% threshold: 0.0008
escitalopram median unadjusted
(0 of 6 pruned genes below threshold)
5% threshold: 0.00083
escitalopram median MDD PGS adj
(0 of 6 pruned genes below threshold)
5% threshold: 0.00082
escitalopram maximum unadjusted
(0 of 6 pruned genes below threshold)
5% threshold: 0.00062
escitalopram maximum MDD PGS adjusted
(0 of 6 pruned genes below threshold)
5% threshold: 0.00063
sertraline median unadjusted
(1 of 13 pruned genes below threshold)
5% threshold: 0.00091
sertraline median MDD PGS adjusted
(1 of 12 pruned genes below threshold)
5% threshold: 0.0009
sertraline maximum unadjusted
(1 of 14 pruned genes below threshold)
5% threshold: 0.00072
sertraline maximum MDD PGS adjusted
(0 of 14 pruned genes below threshold)
5% threshold: 0.00073
Count
P−value

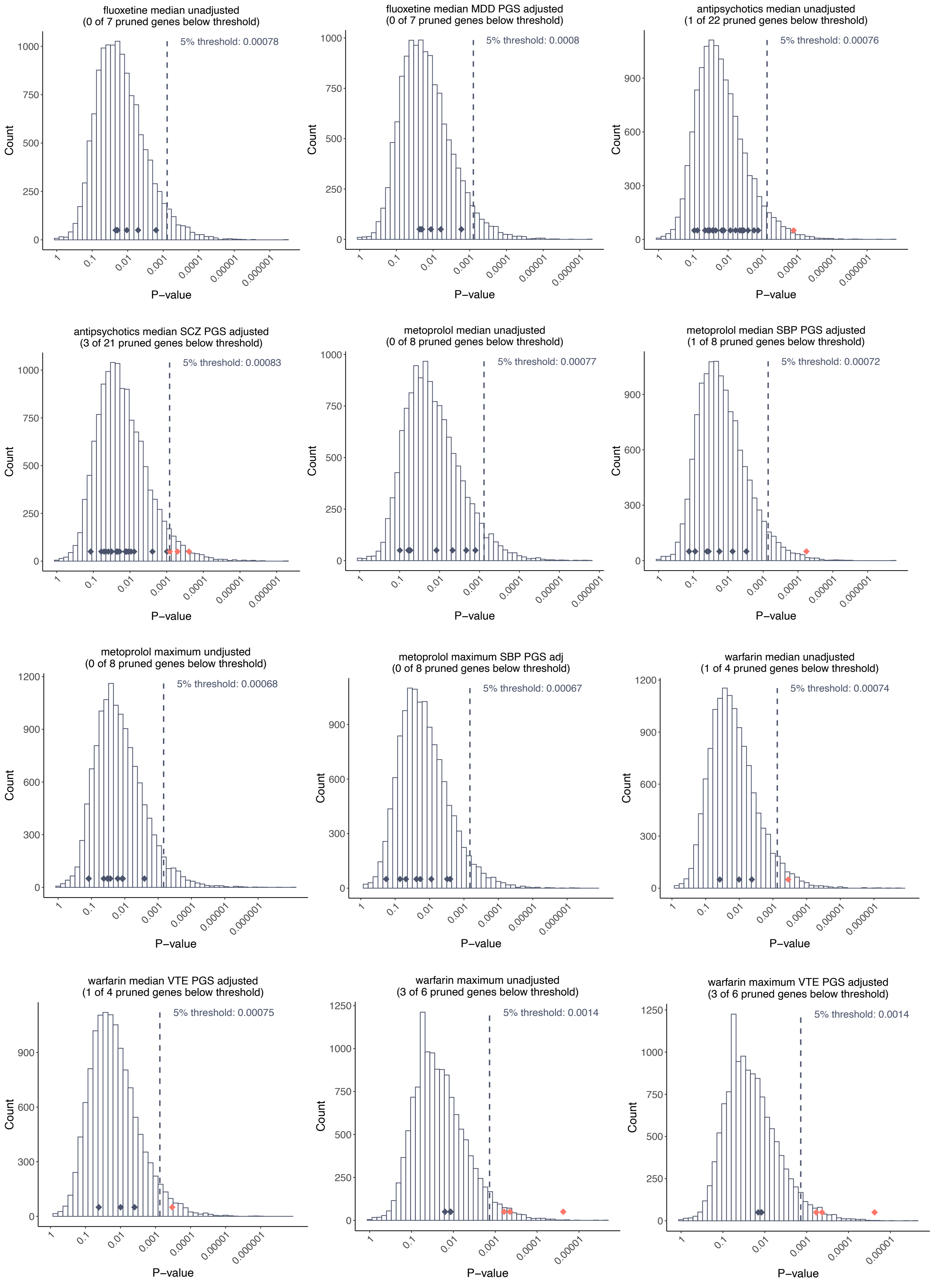

**Supplementary Figure 21. P-value distribution of LD-pruned background genes and PGx genes in GWAS for median and maximum doses of considered drugs.** The histogram provides the distribution of p-values from the background set, ordered from the highest (left) to the lowest (right) p-value on log10 scale. For each gene, the top SNV (i.e., the one with the strongest association) was selected. The dashed vertical line represents the 5th percentile significance threshold, derived from the background set. Dark blue diamonds indicate PGx genes below the threshold, while red diamonds denote PGx genes above the threshold. The number of pruned genes and genes surpassing the threshold are provided in the title. Of note, *CYP2D6* for metoprolol median and maximum dose, and *VKORC1* and *CYP2C9* for warfarin median dose were excluded as top signals for better plot readability, given their strong signals in respective GWAS results. Detailed results, including the top associated SNVs for each PGx gene, are provided in Supplementary Table 8.
